# A Systematic Review of Air Pollution Exposure and Brain Structure and Function during Development

**DOI:** 10.1101/2024.09.13.24313629

**Authors:** Jessica Morrel, Michelle Dong, Michael A. Rosario, Devyn L. Cotter, Katherine L. Bottenhorn, Megan M. Herting

## Abstract

**Objectives:** Air pollutants are known neurotoxicants. In this updated systematic review, we evaluate new evidence since our 2019 systematic review on the effect of outdoor air pollution exposure on childhood and adolescent brain structure and function as measured by magnetic resonance imaging (MRI).

**Methods:** Using PubMed and Web of Science, we conducted an updated literature search and systematic review of articles published through March 2024, using key terms for air pollution and functional and/or structural MRI. Two raters independently screened all articles using Covidence and implemented the risk of bias instrument for systematic reviews informing the World Health Organization Global Air Quality Guidelines.

**Results:** We identified 222 relevant papers, and 14 new studies met our inclusion criteria. Including six studies from our 2019 review, the 20 publications to date include study populations from the United States, Netherlands, Spain, and United Kingdom. Studies investigated exposure periods spanning pregnancy through early adolescence, and estimated air pollutant exposure levels via personal monitoring, geospatial residential estimates, or school courtyard monitors. Brain MRI occurred when children were on average 6-14.7 years old; however, one study assessed newborns. Several MRI modalities were leveraged, including structural morphology, diffusion tensor imaging, restriction spectrum imaging, arterial spin labeling, magnetic resonance spectroscopy, as well as resting-state and task-based functional MRI. Air pollutants were associated with widespread brain differences, although the magnitude and direction of findings are largely inconsistent, making it difficult to draw strong conclusions.

**Conclusion:** Prenatal and childhood exposure to outdoor air pollution is associated with structural and functional brain variations. Compared to our initial 2019 review, publications doubled—an increase that testifies to the importance of this public health issue. Further research is needed to clarify the effects of developmental timing, along with the downstream implications of outdoor air pollution exposure on children’s cognitive and mental health.

**Highlights:**

- Air pollutants have emerged as ubiquitous neurotoxicants
- This field of study has grown substantially in the last 5 years
- Exclusion of highly exposed populations poses a barrier to result generalizability
- Longitudinal investigations are needed to understand developmental impacts of AP
- Additional research should aim to clarify which children may be most vulnerable

Graphical Abstract

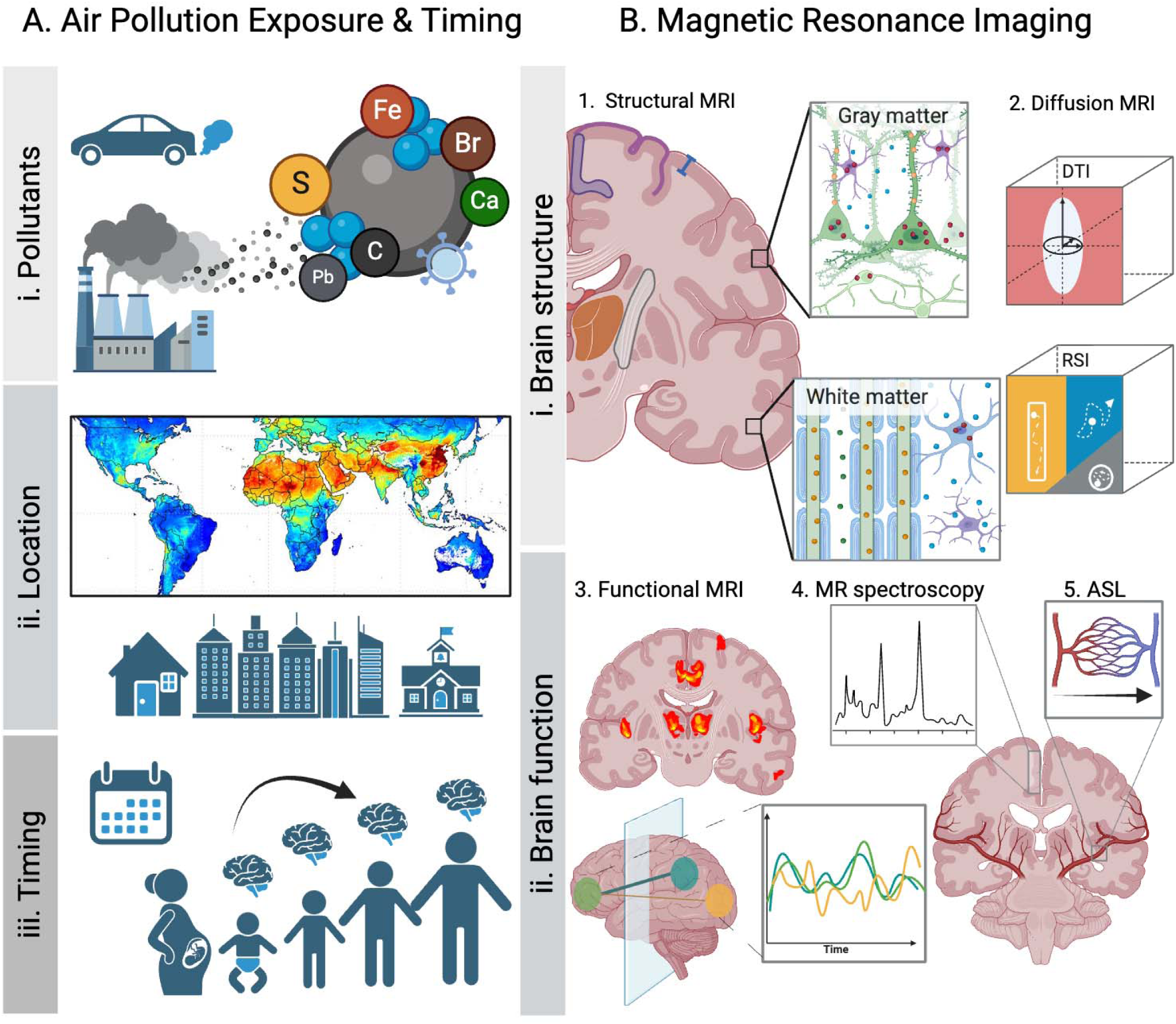

## 1. Introduction

Outdoor ambient air pollution is a hazardous mixture of chemicals and gasses stemming from primary natural (i.e., wildfires, volcanic activity, decomposition of biological material) and anthropogenic (i.e., traffic, power generation, factories) sources, and secondary atmospheric reactions (IARC Working Group on the Evaluation of Carcinogenic Risks to Humans, 2016). Criteria pollutants, for which acceptable standards have been set by the Environmental Protection Agency (EPA), commonly investigated in the human health literature include particulate matter (PM), ground-level ozone (O_3_), and nitrogen dioxide (NO_2_). PM is a mixture of solid particles and liquid droplets suspended in the air and can be categorized depending on size: ultrafine (UFPs; <0.1 μm), fine (PM_2.5_; <2.5 μm), and coarse particles (PM_10_; <10 μm) (US EPA, 2016a). Depending on source(s), various components can be found in PM, including several metals (e.g., lead [Pb], nickel [Ni], iron [Fe]), organic carbon (OC), elemental carbon (EC), sulfate (e.g., SO_4_), and ammonium (NH_4_). These components differ in chemical properties and are produced by different sources; for example, EC stems primarily from diesel engines while NH_4_ is released by fertilizer used in agriculture (Shaltout et al., 2015). Polycyclic aromatic hydrocarbons (PAHs) can also be found in PM; PAHs are chemicals produced through combustion of anthropogenic and natural matter (e.g., coal, gas, and wood; “Toxicological Profile for Polycyclic Aromatic Hydrocarbons,” 1995). In terms of gaseous pollutants, O_3_ is the main pollutant in smog, created by harmful chemical reactions between nitrogen oxides (NO_x_) and volatile organic compounds (US EPA, 2015). NO_2_ is a highly reactive gas that is emitted through fuel combustion (US EPA, 2016b). While overall air quality has been increasing in recent decades (US EPA, 2022), the rapidly worsening climate crisis is changing this landscape. Research suggests that increasing pollution levels may result from climate-induced changes in meteorologic conditions and chemical combustion (Ebi and McGregor, 2008). Thus, given ambient air pollution is ubiquitous and a major environmental health problem, research is imperative to comprehensively understand the potential biological effects of air pollution.

The body is vulnerable to pollutant exposure as early as gestation (for review, see Fussell et al., 2024). Certain pollutants, when inhaled by the mother, have the ability to damage the blood-placenta barrier, while others might even cross it and reach the fetus, leading to bioaccumulation in growing tissues (Bongaerts et al., 2022; Bové et al., 2019; Dong et al., 2018; Wierzba et al., 2018; for review see Bongaerts et al., 2020). Following birth, humans are exposed to air pollution through respiration. Upon inhalation, particles are taken up into the airways through the mouth or nose, where they can be carried to the lungs (Oberdörster et al., 2004; for review see Genc et al., 2012). This invasion can trigger chronic systemic inflammation in the periphery, leading to oxidative stress and cellular damage (Ghio et al., 2012; Rao et al., 2018), which has many downstream physiological and psychological consequences (Hahad et al., 2020). In the lungs, smaller particles can cross the alveoli into the bloodstream where they can be circulated throughout the body (Furuyama et al., 2009; Nemmar et al., 2002). From the blood, some air pollutants are then able to cross the blood-brain barrier (BBB), inducing inflammation and BBB damage (Heidari Nejad et al., 2015; Kang et al., 2021). In the nasal cavity, pollutant particles can contribute to breakdown of the nasal epithelium, where it is hypothesized that they may travel along the olfactory nerve to the olfactory bulb and into the olfactory cortex (Ajmani et al., 2016).

Exposure to air pollution is increasingly implicated as a risk factor for physical and mental health conditions (Elder et al., 2015; Hahad et al., 2020). Emerging evidence suggests that exposure to air pollutants has widespread consequences for brain health across the lifespan, including impairments in general cognitive functioning (Holm et al., 2023; Sakhvidi et al., 2022), neurodevelopmental and mental health conditions such as depression (Borroni et al., 2022; Braithwaite et al., 2019; Yang et al., 2023; Zundel et al., 2022), anxiety (Braithwaite et al., 2019; Yang et al., 2023; Zundel et al., 2022), Attention-Deficit Hyperactivity Disorder (ADHD; Aghaei et al., 2019; Thygesen et al., 2020) and Autism Spectrum Disorder (Flanagan et al., 2023; Lin et al., 2021), as well as increased risk for and/or prevalence of neurodegenerative disease (Calderón-Garcidueñas and Ayala, 2022; Jankowska-Kieltyka et al., 2021; Kuntić et al., 2024; Roy and D’Angiulli, 2024) such as dementia and Alzheimer’s Disease (AD; Hussain et al., 2023; Shi et al., 2020; Zhang et al., 2023), Parkinson’s Disease (Murata et al., 2022; Palacios, 2017; Shi et al., 2020), and Multiple Sclerosis (Farahmandfard et al., 2021; Hedström et al., 2023; Noorimotlagh et al., 2021). While the number of studies exploring these topics is rapidly increasing, the underlying neurobiological mechanisms between pollutant exposure and brain health remain poorly understood. Yet, understanding the etiological pathway(s) from air pollution exposure to adverse cognitive and mental health outcomes has critical implications for the identification of protective and preventive strategies.

While air pollution exposure has been linked to neurological health across the lifespan, the developing brain may be especially susceptible to air pollution neurotoxicity due to rapid and dynamic brain changes that occur from gestation through adolescence (Cardenas-Iniguez et al., 2022; Fox et al., 2010; Herting et al., 2024). The human brain begins developing shortly after conception, with neurogenesis and neuronal migration peaking during the prenatal period (Khodosevich and Sellgren, 2023). Following birth, axonogenesis and synaptogenesis aid in the rapid expansion of white and gray matter volumes, while myelination works in parallel to provide the foundation for brain connectivity and neural propagation (Grotheer et al., 2022). Synaptic pruning begins shortly before the fourth year of life, as these neural connections are selectively eliminated (for review see Khodosevich and Sellgren, 2023). Moreover, neuroimaging advances have allowed researchers to map various brain growth charts. Bethlehem and colleagues (2022) recently charted gray matter, white matter, subcortical, and ventricular volume development using structural Magnetic Resonance Imaging (MRI), showing each neural substrate follows a unique trajectory characterized by differences in velocity and age of peak. For instance, while gray matter volume peaks around mid-childhood, subcortical volume does not reach its peak until early adolescence, and white matter volume continues to increase until young adulthood. Across the cortex, brain regions also reach peak volume at different ages (Bethlehem et al., 2022). By leveraging these neuroimaging methods alongside the existing development MRI literature, MRI studies hold great promise for investigating how air pollution exposure impacts the developing human brain.

For these reasons, in 2019, our team conducted a systematic review to capture the emerging literature exploring associations between early-life air pollution exposure and brain structure and/or function in children and adolescents (Herting et al., 2019). Since this initial review, which identified 6 cross-sectional studies, there has been exponential growth in this area of research. As such, the aim of the current systematic review was to provide an updated synthesis of the literature exploring structural and/or functional MRI brain correlates of outdoor air pollution exposure in children and adolescents. This review uses a PRISMA (Page et al., 2021) framework to identify relevant articles meeting inclusion criteria published through March 2024. Risk of bias was assessed using a modified version of the World Health Organization (WHO) Risk of Bias Instrument for Systematic Reviews Informing WHO Global Air Quality Guidelines (WHO Global Air Quality Guidelines Working Group on Risk of Bias Assessment, 2020).

## 2. Materials and Methods

### 2.1 Search Strategy

Extensive searches of PubMed and Web of Science were performed to identify studies examining relationships between outdoor air pollution exposures and MRI brain outcomes in children and adolescents. In this updated systematic review, we identified articles published since the initial review conducted from our team (i.e., after December 5, 2019). To identify these relevant studies, our team used the exact search strategy outlined previously (Herting et al., 2019), including MeSH terms for air pollution and functional and/or structural MRI, with inclusion criteria of English-language, human species, primary research articles, and children and/or adolescents (<24 years-old). The following search algorithm was used: [(“magnetic resonance image”[title/abstract] OR “magnetic resonance images”[title/abstract] OR “magnetic resonance imaging”[title/abstract] OR “MRI” [Title/Abstract] OR “white matter hyperintensity”[title/abstract] OR “white matter hyperintensities”[title/abstract] OR “neuroimage”[title/abstract] OR “neuroimages”[title/abstract] OR “neuroimaging”[title/abstract] OR “neuroinflammation”[title/abstract] OR “systemic inflammation”[title/abstract] OR “white matter volume”[title/abstract] OR “white matter volumes”[title/abstract] OR “brain structure”[title/abstract] OR “brain volume”[title/abstract] OR “brain volumes”[title/abstract] OR “neurotoxic”[Title/Abstract] OR “neurotoxicity”[Title/Abstract] OR “neurotoxicities”[Title/Abstract] OR “functional connectivity”[Title/Abstract] OR “Brain/pathology”[mesh] OR “Brain/physiopathology”[Majr] OR “Magnetic Resonance Imaging”[Mesh] OR “Cognition Disorders/pathology”[Mesh] OR “Cognition Disorders/chemically induced”[Majr] OR “White Matter/pathology”[Majr]) AND (“Air Pollution”[Title/Abstract] OR “Air Pollutant”[Title/Abstract] OR “Air Pollutants”[Title/Abstract] OR “Particulate Matter”[Title/Abstract] OR “Ozone”[Title/Abstract] OR “Nitrogen dioxide”[Title/Abstract] OR “Nitrogen oxides”[Title/Abstract] OR “Sulfur Dioxide”[Title/Abstract] OR “black carbon”[title/abstract] OR “elemental carbon”[title/abstract] OR “Vehicle Emission”[Title/Abstract] OR “Vehicle Emissions”[Title/Abstract] OR “diesel”[Title/Abstract] OR “diesel exhaust”[Title/Abstract] OR “diesel exhausts”[Title/Abstract] OR “vehicle exhaust”[Title/Abstract] OR “vehicle exhausts”[Title/Abstract] OR “vehicular exhaust”[Title/Abstract] OR “vehicular exhausts”[Title/Abstract] OR “road traffic”[Title/Abstract] OR “PM2.5”[Title/Abstract] OR “PM10”[Title/Abstract] OR “coarse particle”[Title/Abstract] OR “coarse particles”[Title/Abstract] OR “ultrafine particle”[Title/Abstract] OR “ultrafine particles”[Title/Abstract] OR “Polycyclic aromatic hydrocarbon”[Title/Abstract] OR “Polycyclic aromatic hydrocarbons”[Title/Abstract] OR “Air Pollution”[Mesh] OR “Particulate Matter”[Mesh] OR “Ozone”[Mesh] OR “Nitrogen dioxide”[Mesh] OR “Nitrogen oxides”[Mesh] OR “Sulfur Dioxide”[Mesh] OR “Vehicle Emissions”[Mesh] OR “Air Pollution/adverse effects”[Majr] OR “Polycyclic Aromatic Hydrocarbons/adverse effects”[Mesh] OR “Polycyclic Aromatic Hydrocarbons/poisoning”[Mesh] OR “Polycyclic Aromatic Hydrocarbons/toxicity”[Mesh] OR “Inhalation Exposure/adverse effects”[Mesh])] (see **Supplemental Table 1** for list of search terms). However, relying solely on MeSH terms for some key concepts inadvertently excluded studies published in journals that do not provide MeSH terms (DeMars and Perruso, 2022). Thus, we conducted a secondary search aimed to identify all articles published any time before March 12, 2024 using key search terms via relevant title/abstract text (i.e., an identical search without MeSH terms and related constraints). The exact search algorithm for this search was: [(“child” OR “adolescent”) AND (”magnetic resonance image”[title/abstract] OR “magnetic resonance images”[title/abstract] OR “magnetic resonance imaging”[title/abstract] OR “MRI” [Title/Abstract] OR “white matter hyperintensity”[title/abstract] OR “white matter hyperintensities”[title/abstract] OR “neuroimage”[title/abstract] OR “neuroimages”[title/abstract] OR “neuroimaging”[title/abstract] OR “neuroinflammation”[title/abstract] OR “systemic inflammation”[title/abstract] OR “white matter volume”[title/abstract] OR “white matter volumes”[title/abstract] OR “brain structure”[title/abstract] OR “brain volume”[title/abstract] OR “brain volumes”[title/abstract] OR “neurotoxic”[Title/Abstract] OR “neurotoxicity”[Title/Abstract] OR “neurotoxicities”[Title/Abstract] OR “functional connectivity”[Title/Abstract]) AND (”Air Pollution”[Title/Abstract] OR “Air Pollutant”[Title/Abstract] OR “Air Pollutants”[Title/Abstract] OR “Particulate Matter”[Title/Abstract] OR “Ozone”[Title/Abstract] OR “Nitrogen dioxide”[Title/Abstract] OR “Nitrogen oxides”[Title/Abstract] OR “Sulfur Dioxide”[Title/Abstract] OR “black carbon”[title/abstract] OR “elemental carbon”[title/abstract] OR “Vehicle Emission”[Title/Abstract] OR “Vehicle Emissions”[Title/Abstract] OR “diesel”[Title/Abstract] OR “diesel exhaust”[Title/Abstract] OR “diesel exhausts”[Title/Abstract] OR “vehicle exhaust”[Title/Abstract] OR “vehicle exhausts”[Title/Abstract] OR “vehicular exhaust”[Title/Abstract] OR “vehicular exhausts”[Title/Abstract] OR “road traffic”[Title/Abstract] OR “PM2.5”[Title/Abstract] OR “PM10”[Title/Abstract] OR “coarse particle”[Title/Abstract] OR “coarse particles”[Title/Abstract] OR “ultrafine particle”[Title/Abstract] OR “ultrafine particles”[Title/Abstract] OR “Polycyclic aromatic hydrocarbon”[Title/Abstract] OR “Polycyclic aromatic hydrocarbons”[Title/Abstract]) AND brain NOT (Review [Publication Type] OR Preprint [Publication Type]). Thus, the combined search strategy included all relevant articles published on Pubmed or Web of Science through March 12, 2024.

### 2.2 Selection Process and Review

**Figure 1** outlines the study selection process below. Unique articles were identified by MeSH terms and text fields. Titles and abstracts of resulting articles were obtained and uploaded to Covidence (Veritas Health Innovation, 2024). The selection process to identify relevant papers was two-fold, including title and abstract screening followed by full-text screening. Two reviewers (M.D. and J.M.) independently screened all abstracts, and a third reviewer (M.A.R.) was consulted to resolve disagreements. After narrowing down relevant papers based on abstracts, studies were selected for full-text review. Additional inclusionary criteria included: (1) full-length original research article, (2) individual-level outdoor air pollution data from a period from *in utero* to time of MRI assessment, (3) brain MRI outcome (e.g., structural, diffusion, functional, arterial spin labeling, spectroscopy), and (4) population of children or adolescents (<24 years; Sawyer et al., 2018). Studies were excluded if they met the following criteria: (1) not an original research paper (e.g., meta-analysis, review paper, or findings published previously), (2) measurements of only indoor or occupational air pollution, (3) community-level exposure estimates, (4) no brain MRI outcome of interest, and (5) studies conducted in individuals aged >24 years.

**Figure 1.**
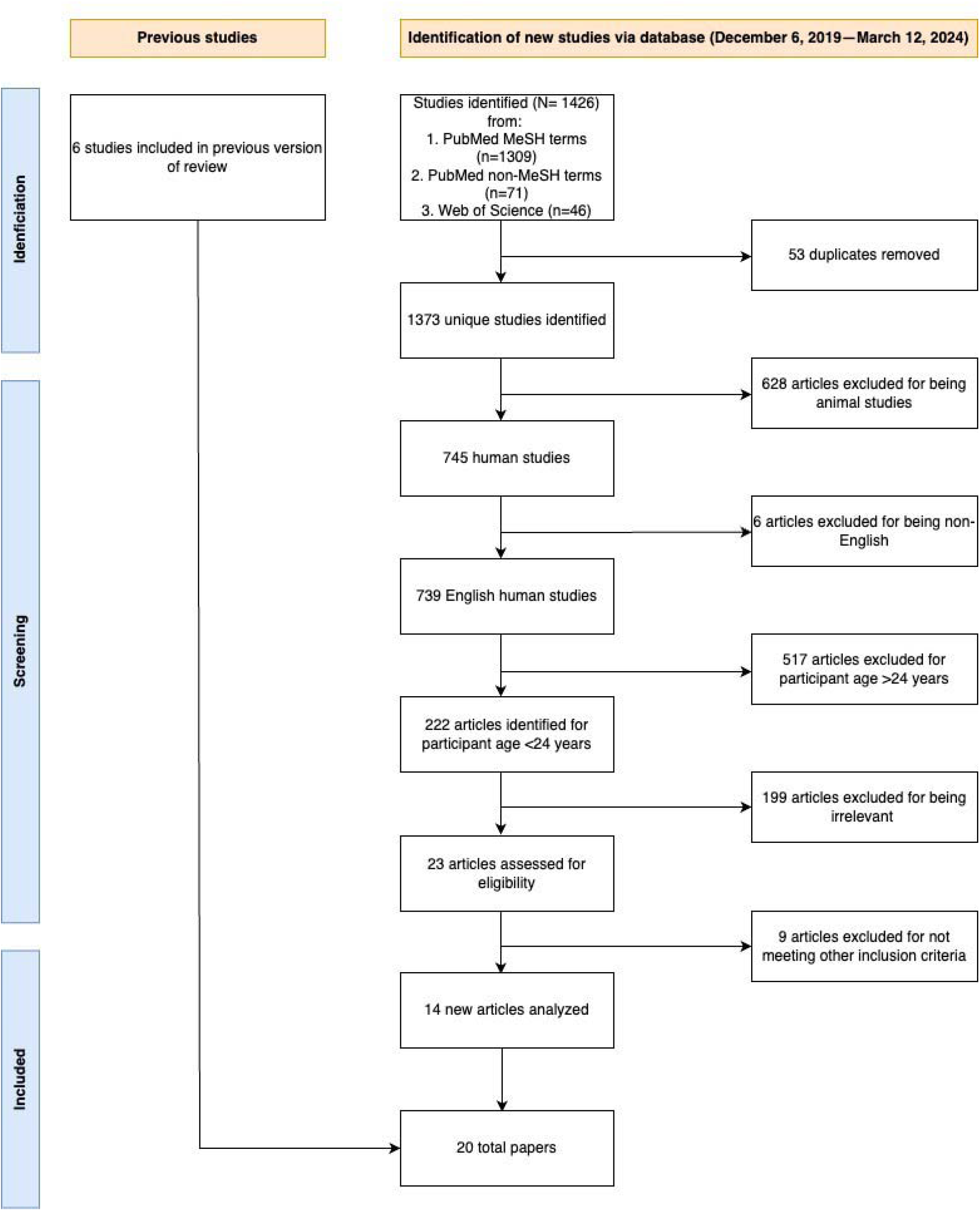
PRISMA flow diagram of study selection for review focused on air pollution and brain MRI in children and adolescents.

### 2.3 Extraction and Risk of Bias

#### 2.3.1 Extraction

The following data were extracted: author, date of publication, study sample/cohort, study design, participant characteristics, pollutant type, timing of exposure, outcome measurement methods, main findings (prenatal and postnatal), null findings, and covariates.

#### 2.3.2 Assessment of Bias

The quality of studies was assessed using a modified risk of bias assessment instrument for systematic reviews based on the WHO Global Air Quality Guidelines (2020). This scale consists of three categories (‘low-risk’, ‘moderate-risk’, and ‘high-risk’) to judge six domains, including: 1) confounding variables, 2) selection bias, 3) exposure assessment, 4) outcome measurement, 5) missing data, and 6) selective reporting. For each domain, related subdomain questions are provided to assist in making a judgment about whether the study presents as ‘low’, ‘moderate’, or ‘high’ risk of bias. If one of the subdomains has a rating of high risk of bias, the whole domain is rated as high risk of bias. The whole domain is rated as low risk of bias only if all the subdomains have a rating of low risk of bias. When one subdomain has a rating of moderate risk of bias and none of the other subdomains are high risk of bias, the whole domain is rated as moderate risk of bias. For the first domain, we chose critical and potential confounding variables to evaluate in each study using *a priori* causal knowledge diagram (Hernán et al., 2002) to identify confounders, defined as variables known to predict the outcome of interest (i.e., brain maturation) and likely to influence exposure to ambient air pollution (**Supplemental Figure 1**). We considered the participants’ age at MRI, sex, and family-level socioeconomic status (SES; i.e., income, parental education) as critical confounders, per WHO guidelines; potential confounders to consider varied depending on study design, but included race and/or ethnicity and/or parental country of birth, other maternal and family-level SES factors, pregnancy related factors, home environment, participant engagement with outdoor environment, other environmental co-exposures, as well as important MRI covariates. Using these criteria, two reviewers (M.D. and J.M.) independently assessed each study, judging based on each of the six domains. After reviewing each study, a third reviewer (M.A.R.) resolved disagreements and synthesized results from the initial review of bias.

## 3. Results

Using the comprehensive search strategy, 1309 articles were identified from PubMed using MeSH terms, and 117 articles were identified using non-MeSH terms. Out of the 1373 unique studies identified, 745 studies met the criteria of only including humans. Six articles were excluded for being in a language other than English, and 517 studies were removed for including participants >24 years of age. Of the remaining 222 articles, 199 were excluded following title and abstract screening, since they did not meet the above criteria. The remaining 23 articles were selected for full-text review, with nine articles excluded for not meeting inclusion criteria. The remaining 14 articles were considered relevant original research articles and were included in the final review (**Figure 1**). Including the six studies reviewed in our team’s 2019 paper, a total of 20 articles represents the current state of the literature examining relationships between air pollution exposure and MRI brain outcomes. Details of the 20 studies are presented in **Table 1**.

**Table 1.**
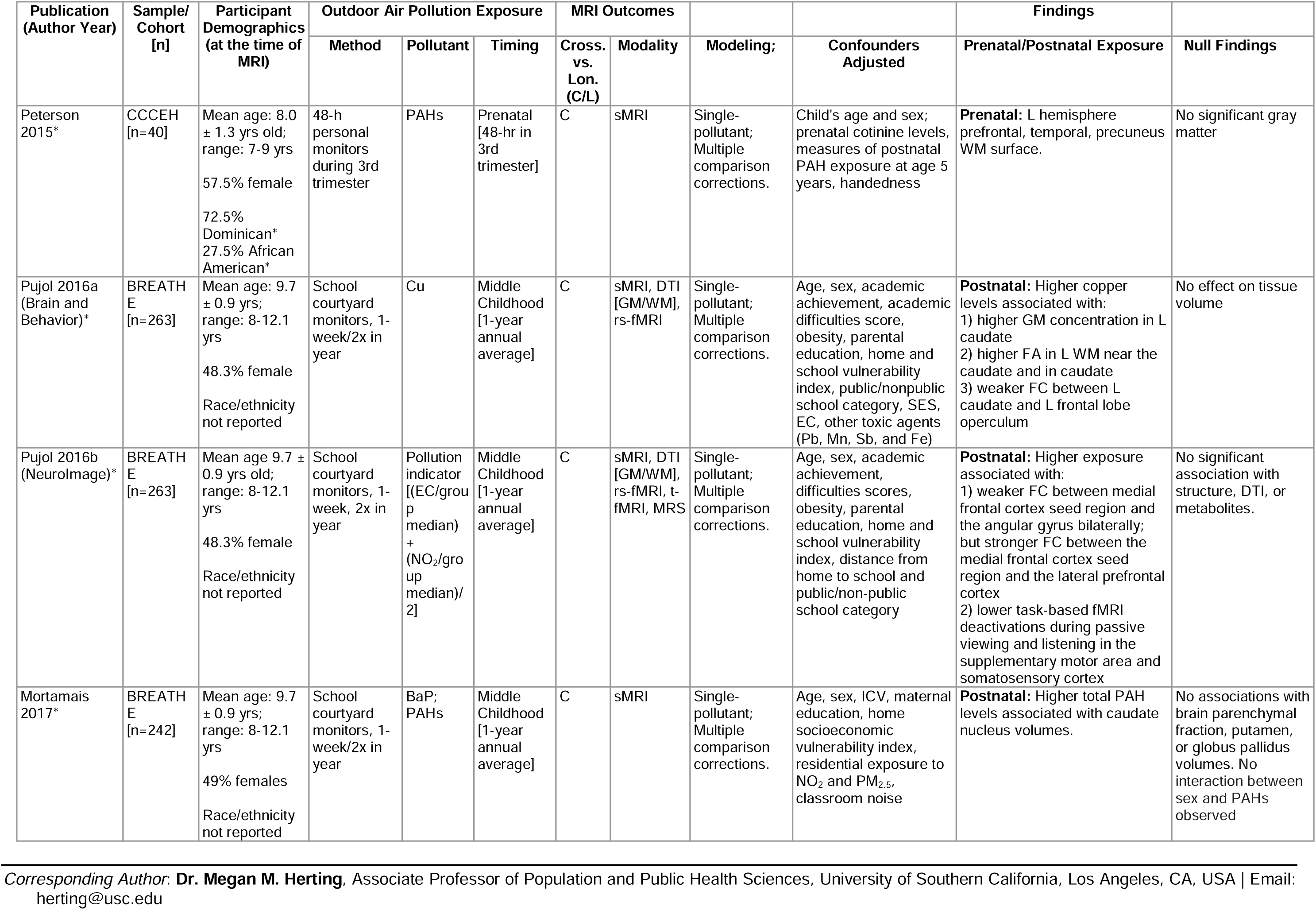
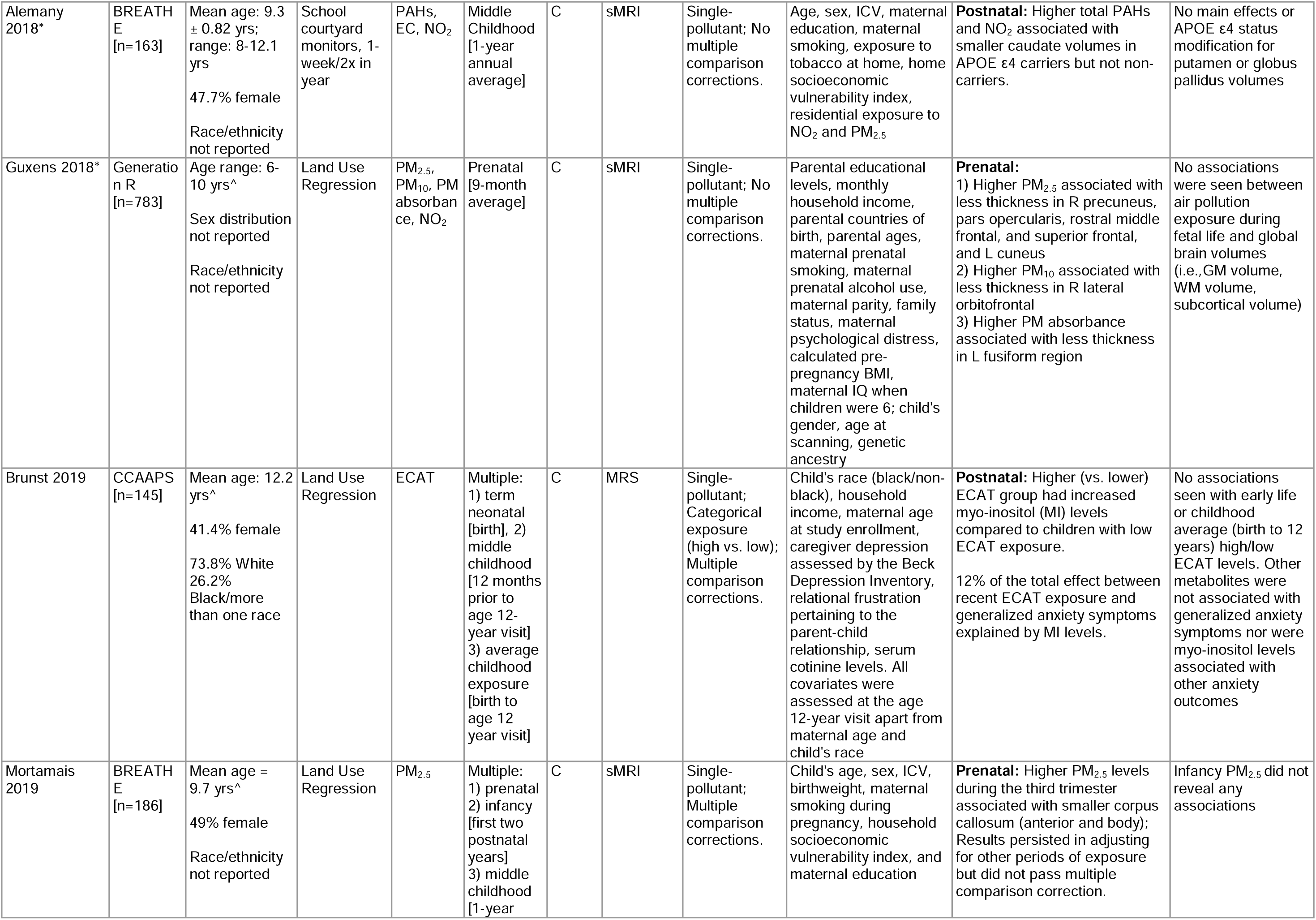
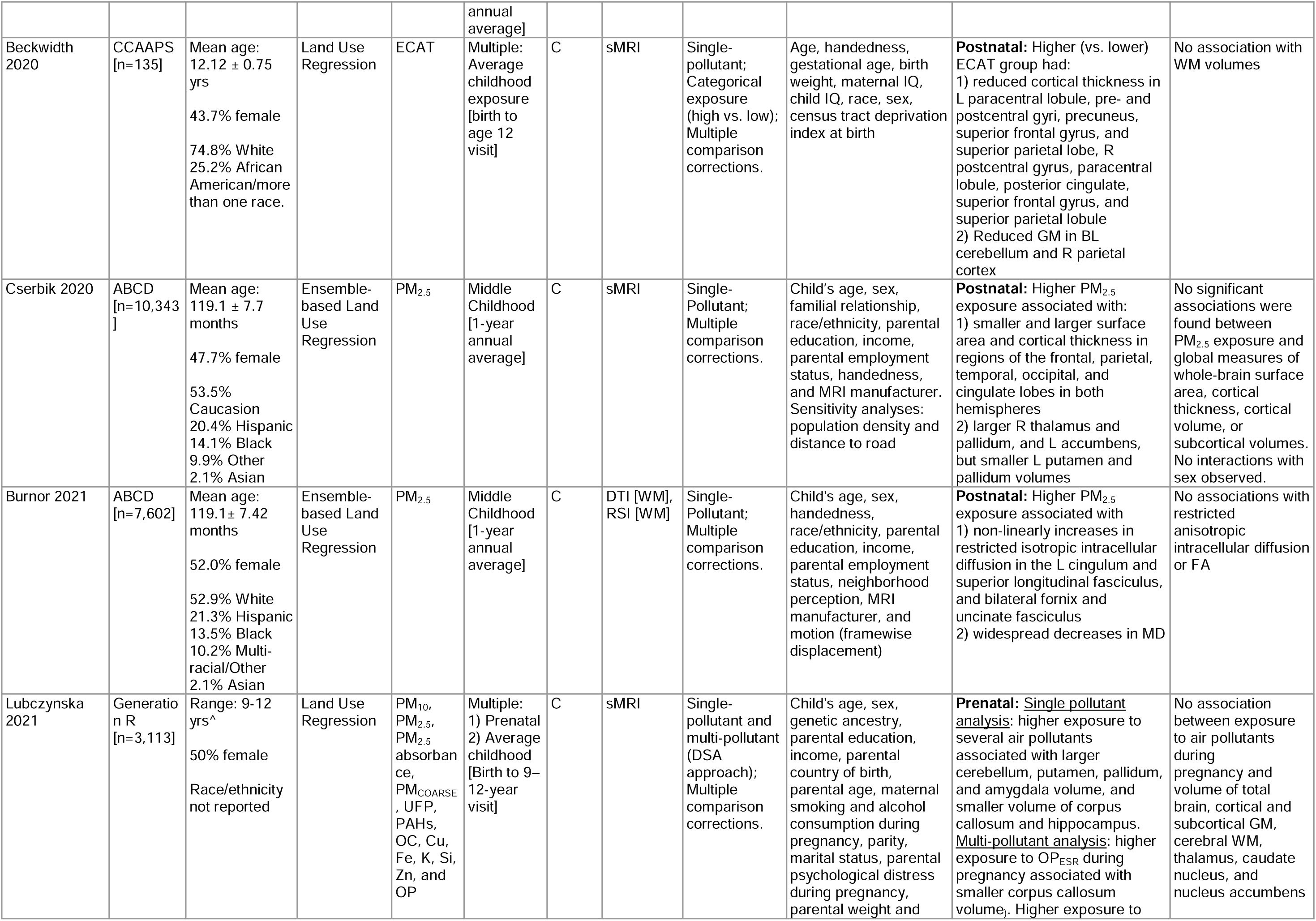
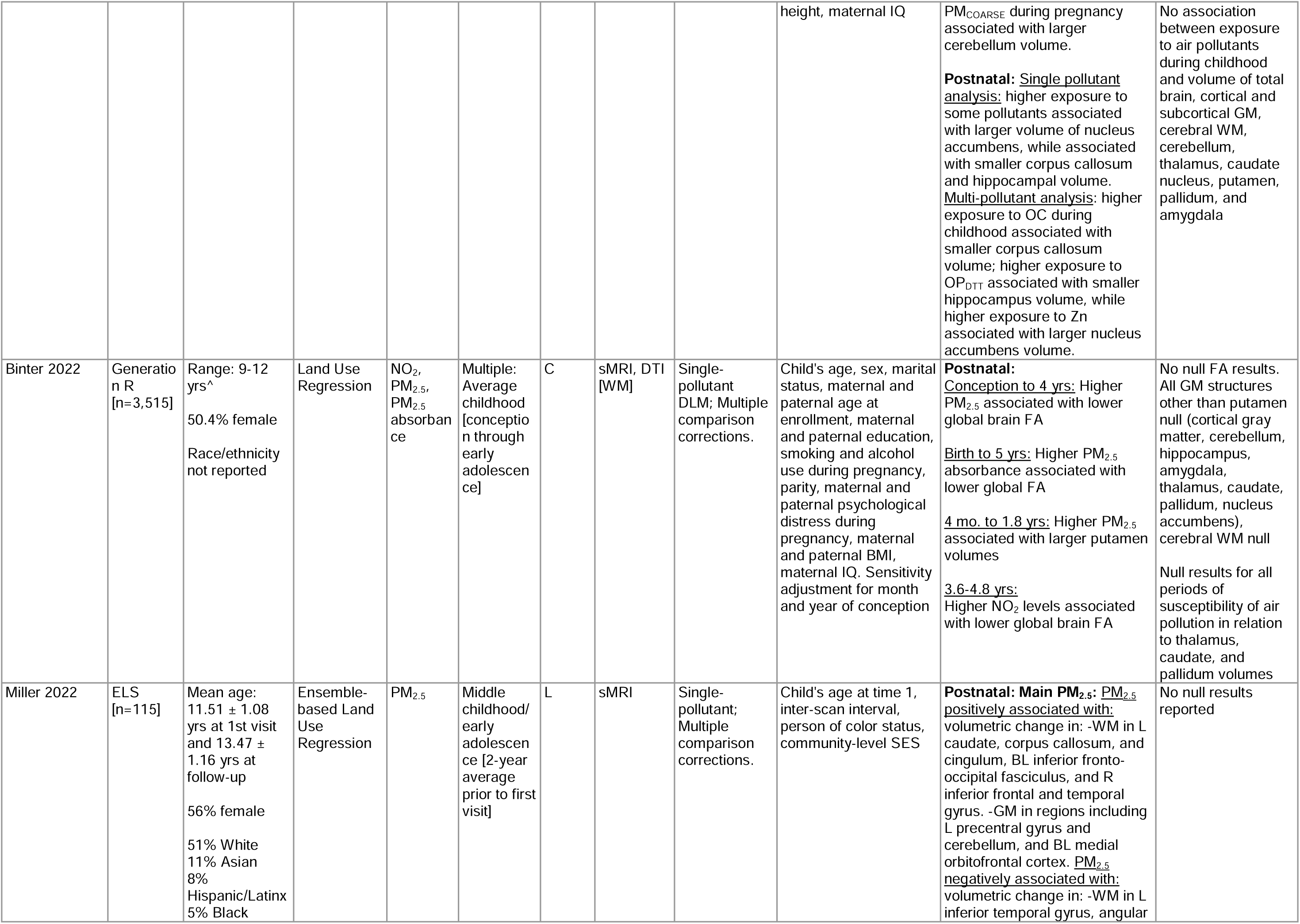
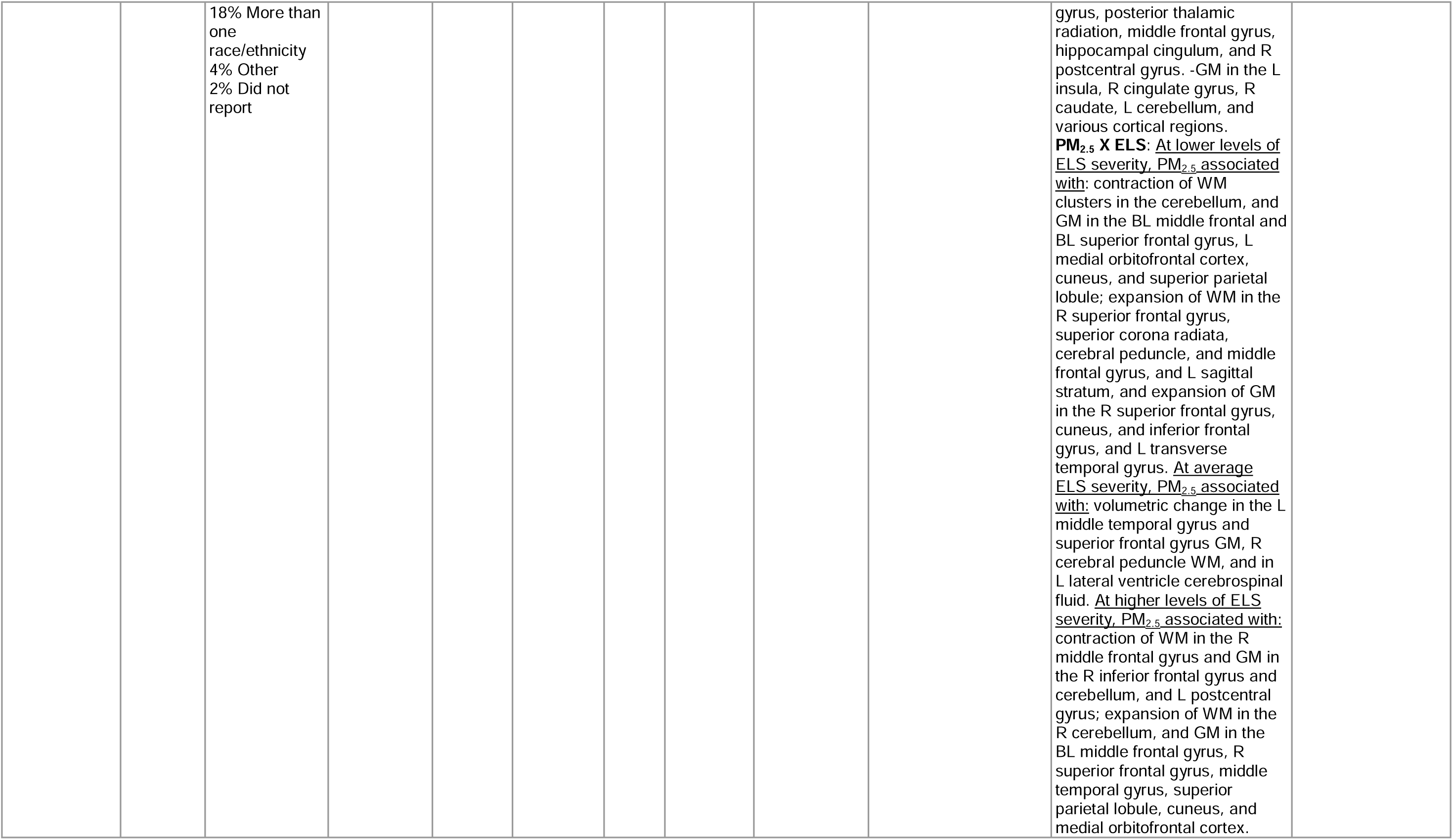
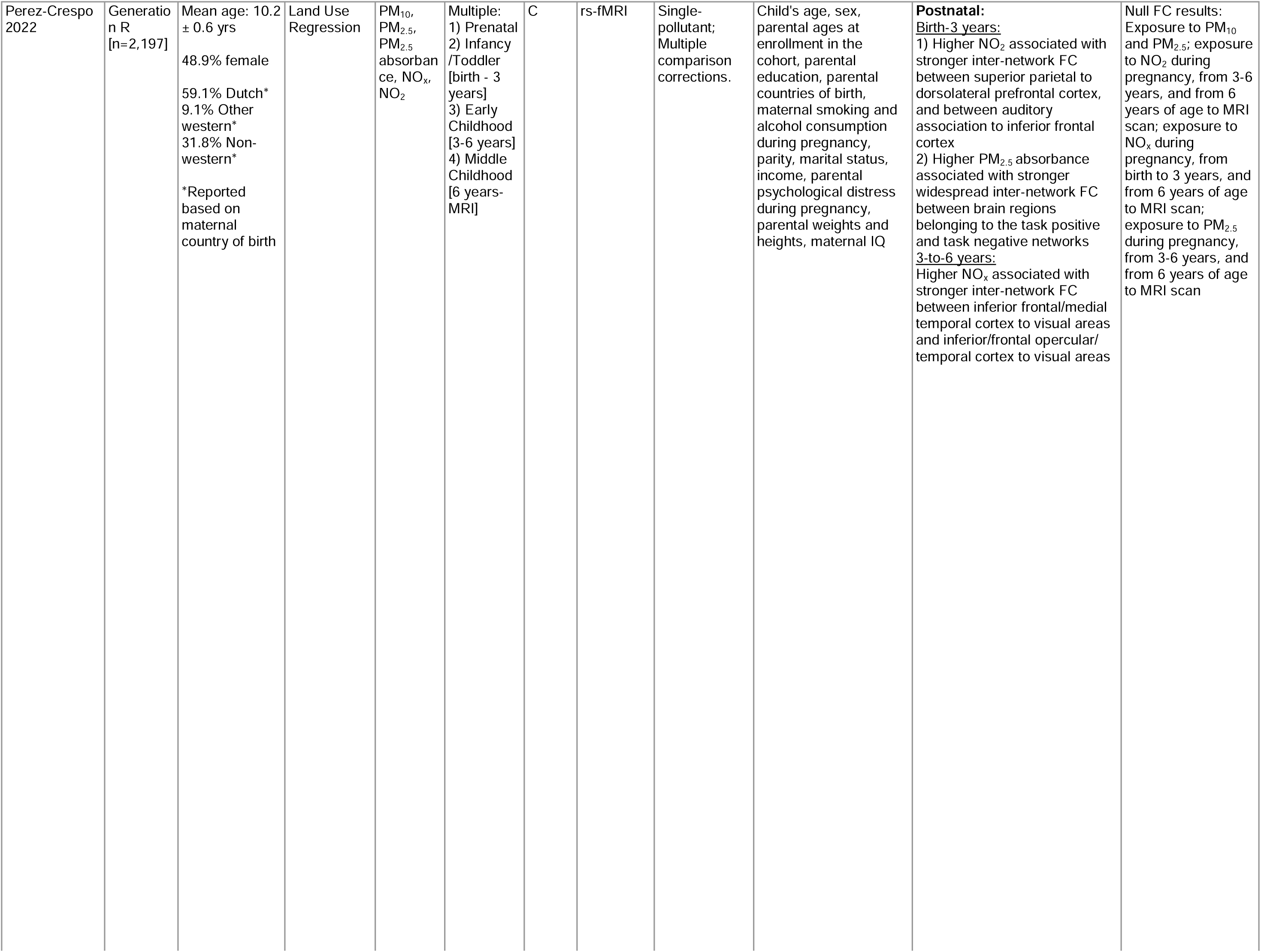
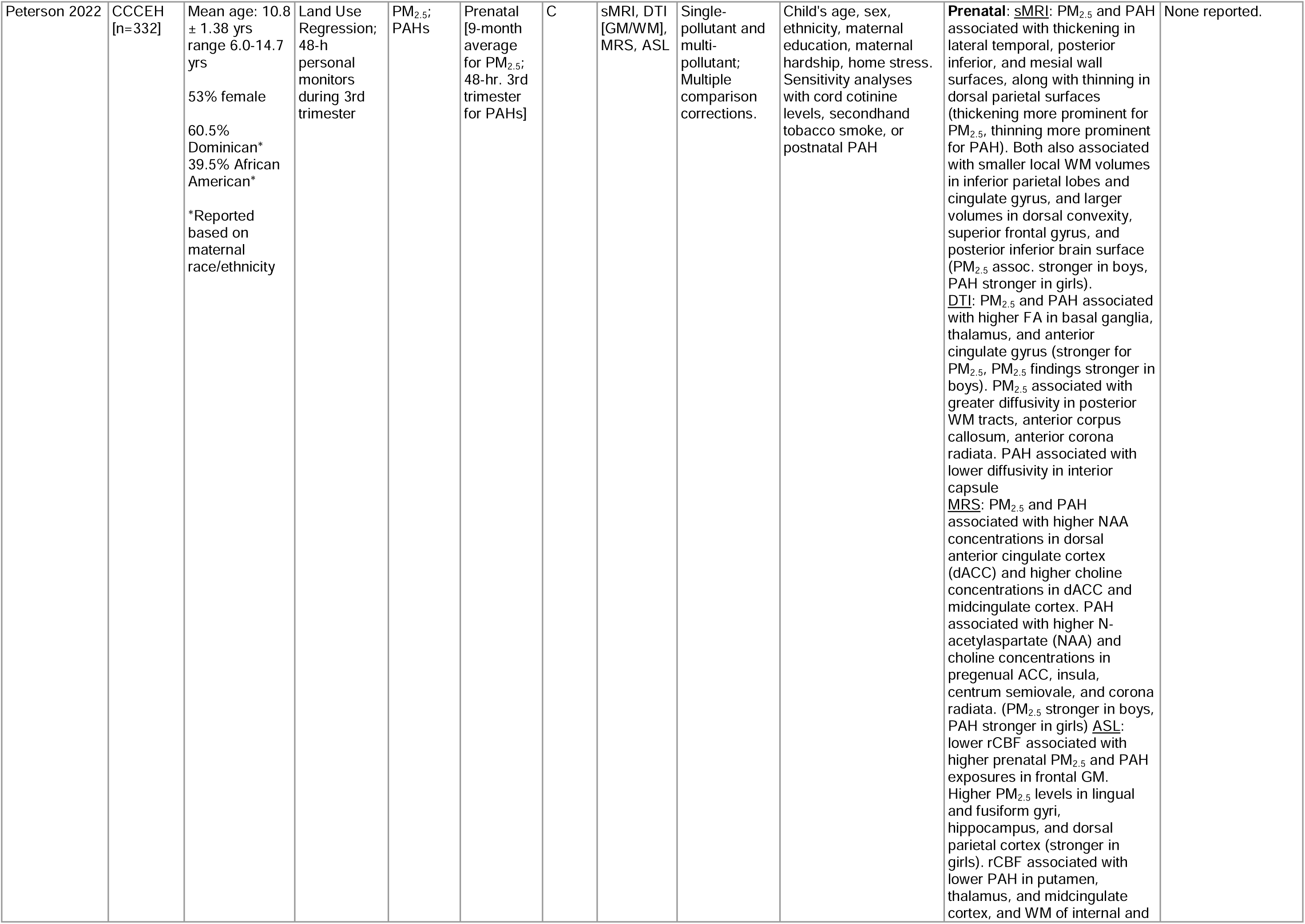
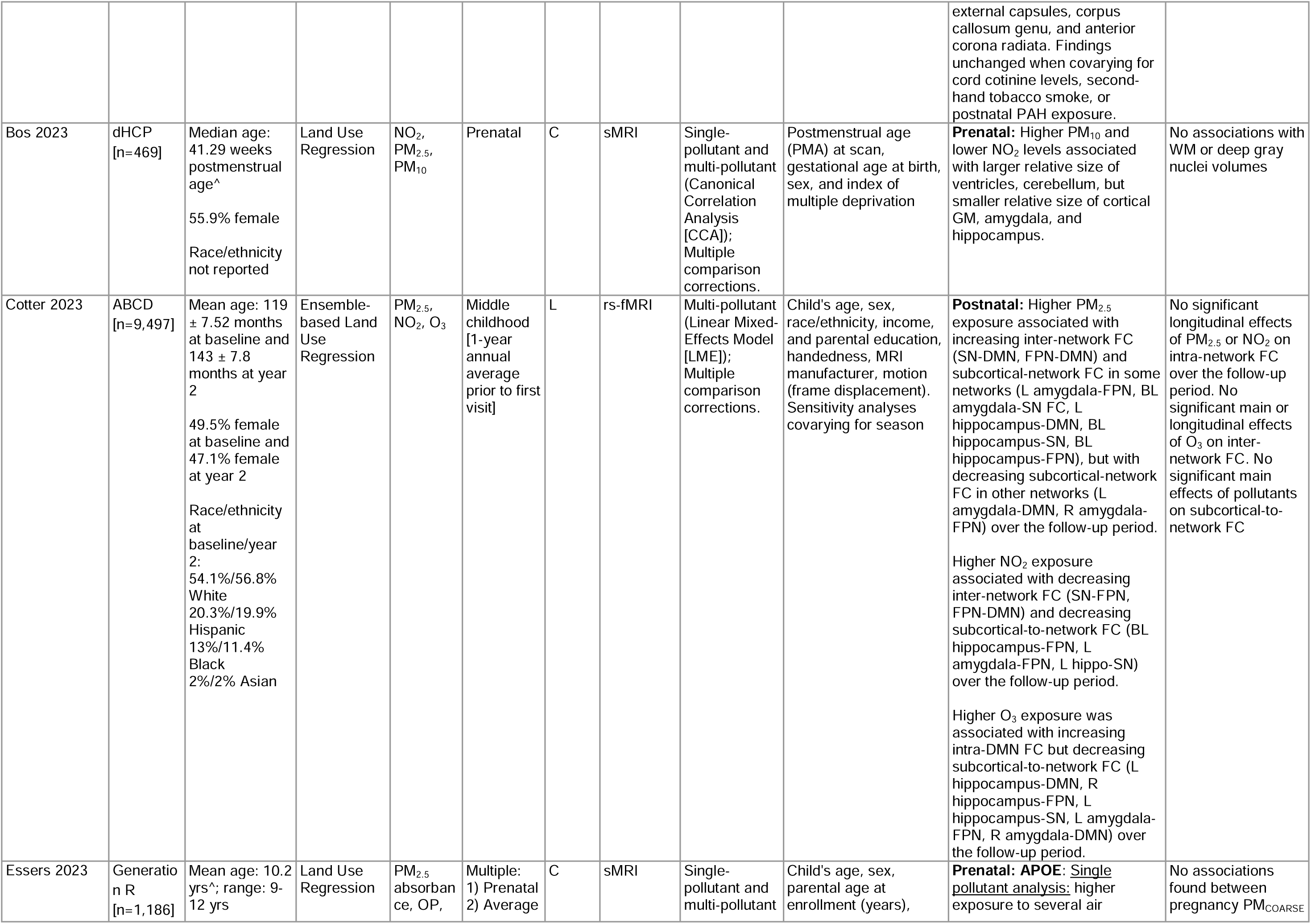
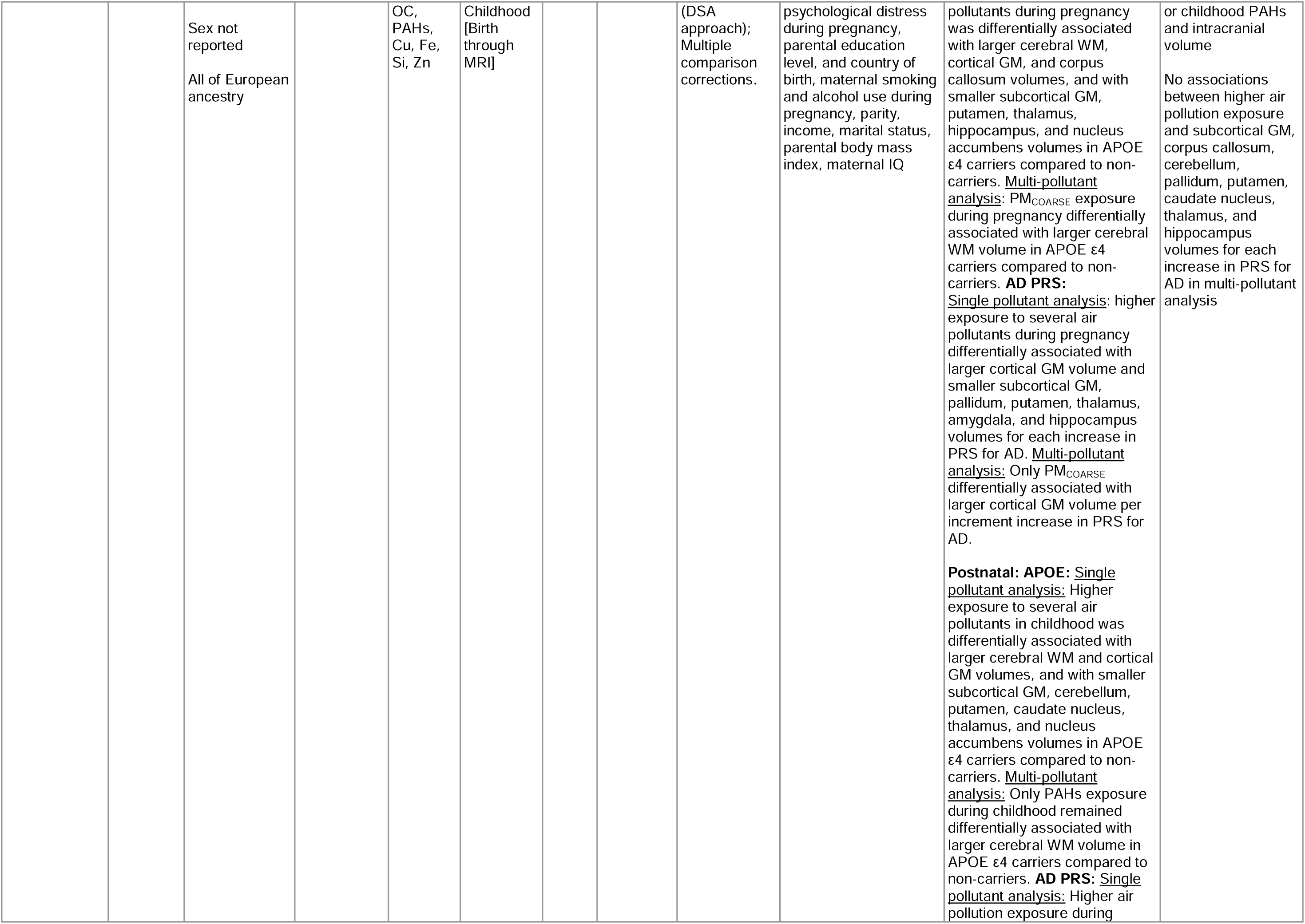
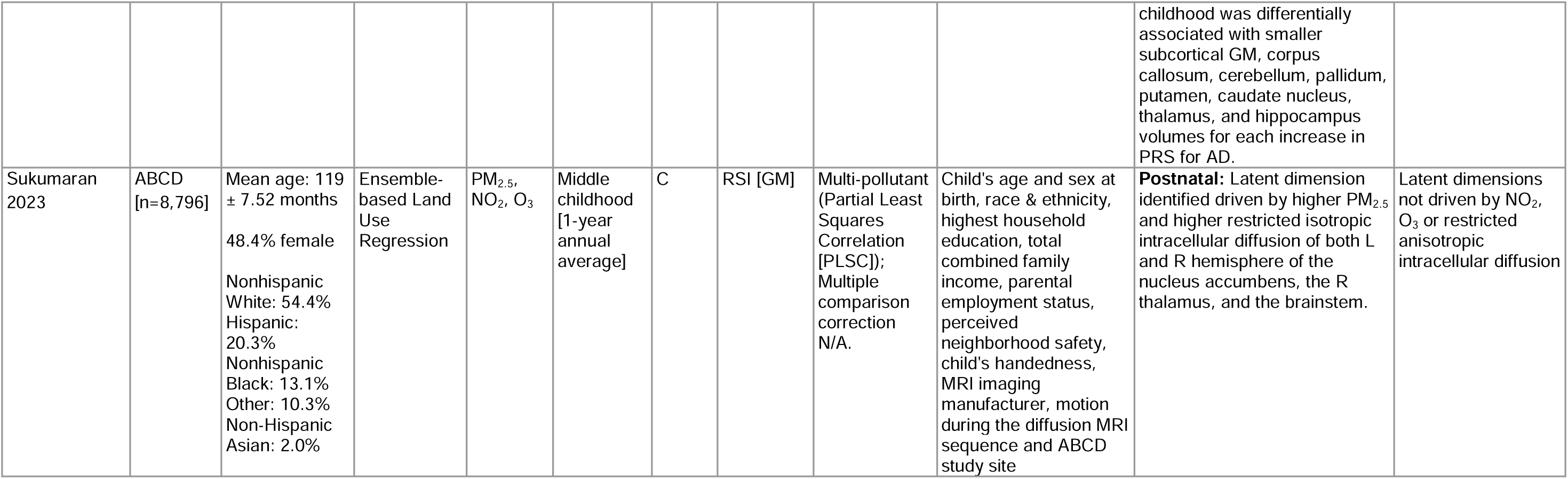
Overview of air pollution exposures, MRI outcomes, significant, and null results from all included studies. Abbreviations: ABCD = Adolescent Brain Cognitive Development Study; BREATHE = B**r**ain Development and Air Pollution Ultrafine Particles in School Children Project; CCAAPS = Cincinnati Childhood Allergy and Air Pollution Study; CCCEH = Columbia Center for Children’s Environmental Health Cohort; dHCP = Developing Human Connectome Project; ELS = Early Life Stress on Neurodevelopment Study; Generation R = Generation R Study. BaP = Benzo[a]pyrene; BC = Black Carbon; Cu = Copper; EC = Elemental Carbon; ECAT = Elemental Carbon Attributable to Traffic; Fe = Iron; K = Potassium; NO_2_ = Nitrogen Dioxide; NO_x_ = Nitric Oxide; O_3_ = Ozone; OC = Organic Carbon; OP = Oxidative Potential of PM_2.5_; OP_ESR_ = OP evaluated using electron spin resonance; OP_DTT_ = OP evaluated using dithiothreitol; PAH = Polycyclic Aromatic Hydrocarbon; PM_2.5_ = Fine Particulate Matter; PM_10_ = Coarse Particulate Matter; PM_COARSE_ = Particulate matter between 2.5-10µm; Si = Silicon; UFP = Ultrafine Particulate Matter; Zn = Zinc. ASL = Arterial Spin Labeling; DTI = Diffusion Tensor Imaging; DWI = Diffusion Weighted Imaging; MRI = Magnetic Resonance Imaging; MRS = Magnetic Resonance Spectroscopy; rs-fMRI= Resting State Functional Magnetic Resonance Imaging; RSI = Restriction Spectrum Imaging; sMRI: Structural Magnetic Resonance Imaging. MD = Mean Diffusivity; FA = Fractional Anisotropy; FC = Functional Connectivity; GM = Gray Matter; ICV = Intracranial volume; rCBF = relative Cerebral Blood Flow; WM = White Matter. BL = Bilateral; DMN = Default Mode Network; FPN = Frontoparietal Network; L = Left; R = Right; SN = Salience Network. DSA = Deletion/Substitution/Addition. BMI = Body Mass Index; SES = Socioeconomic Status; IQ = Intelligence Quotient; AD = Alzheimer’s Disease; APOE = Apolipoprotein E; PRS = polygenic risk score. *Denotes included in previous review. ^Mean and/or standard deviation not reported.

### 3.1.1 Description of cohorts and study participants

Notably, the 20 published studies are based on seven study participant samples, most of which are based on larger birth or child cohorts, and all of which have come from Western countries (**Table 2**; **Figure 2**). We outline the details of each of study samples alphabetically below.

**Figure 2.**
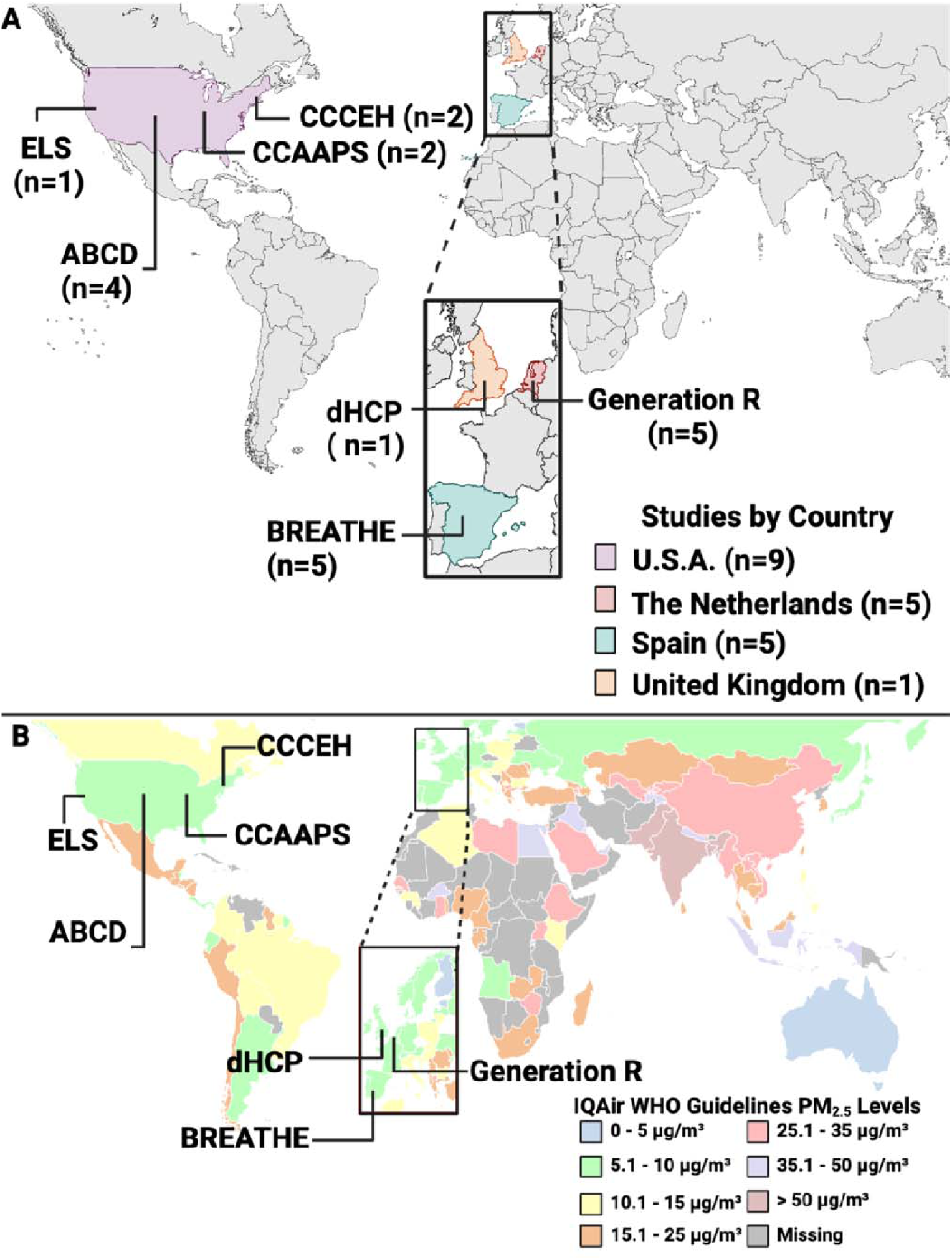
A) Map of study locations for air pollution and brain MRI findings in children most and adolescents (n=20 studies). Cohort and study sample details can be found in **Table 2**. **B) Map of** PM_2.5_ exposure levels worldwide based on IQAir World Health Organization 2023 data. **Most** included studies have come from countries that currently have relatively low levels of exposure to air pollution; albeit exposure levels for any given study may be higher given some studies examined exposure during pregnancy as far back as 1998 when air pollution levels were markedly higher in Western countries. Abbreviations: ABCD = Adolescent Brain Cognitive Development Study; BREATHE = B**r**ain Development and Air Pollution Ultrafine Particles in School Children Project; CCAAPS = Cincinnati Childhood Allergy and Air Pollution Study; CCCEH = Columbia Center for Children’s Environmental Health Cohort; dHCP = Developing Human Connectome Project; ELS = Early Life Stress on Neurodevelopment Study; Generation R = Generation R Study; WHO = World Health Organization. Created with Biorender.

**Table 2.**
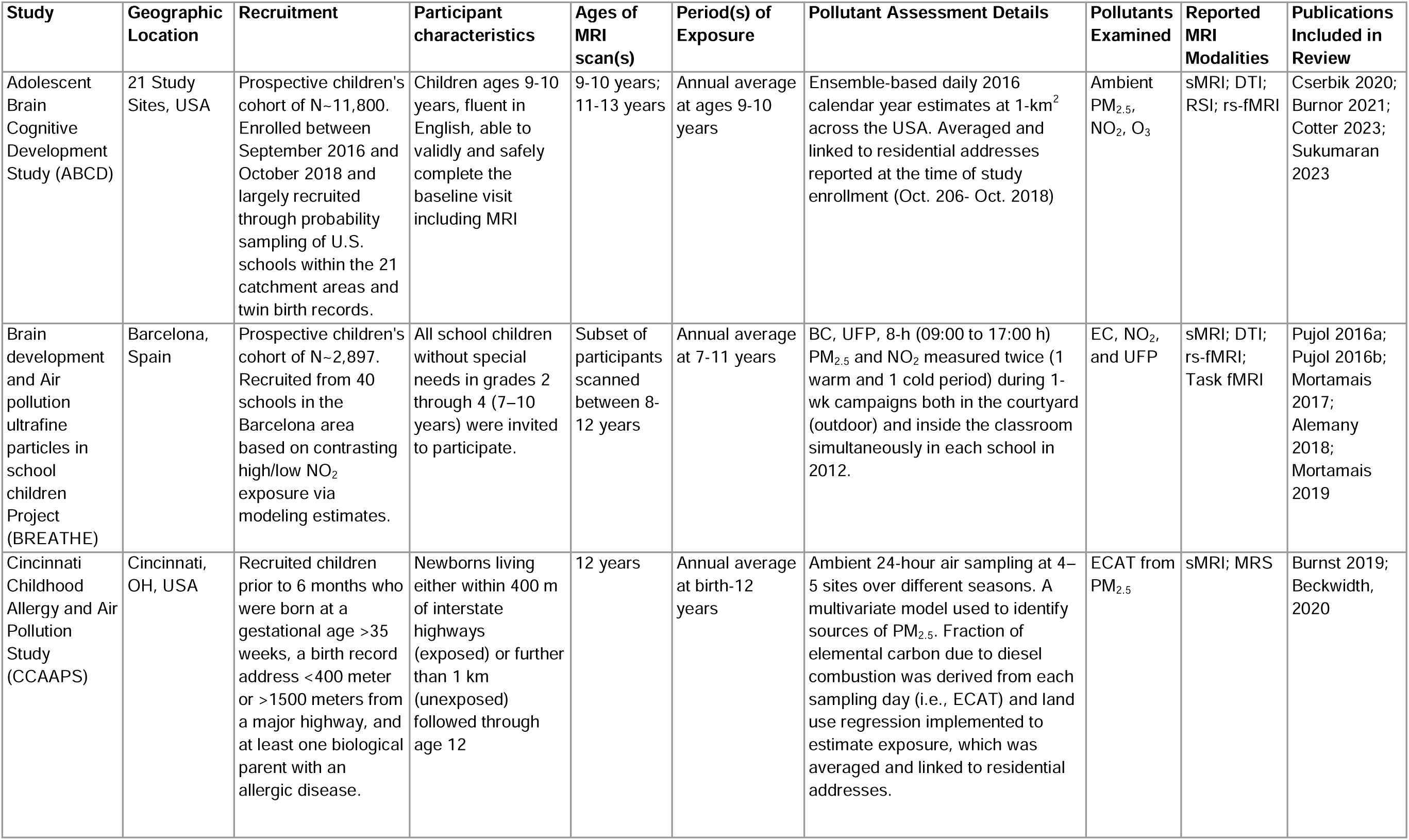
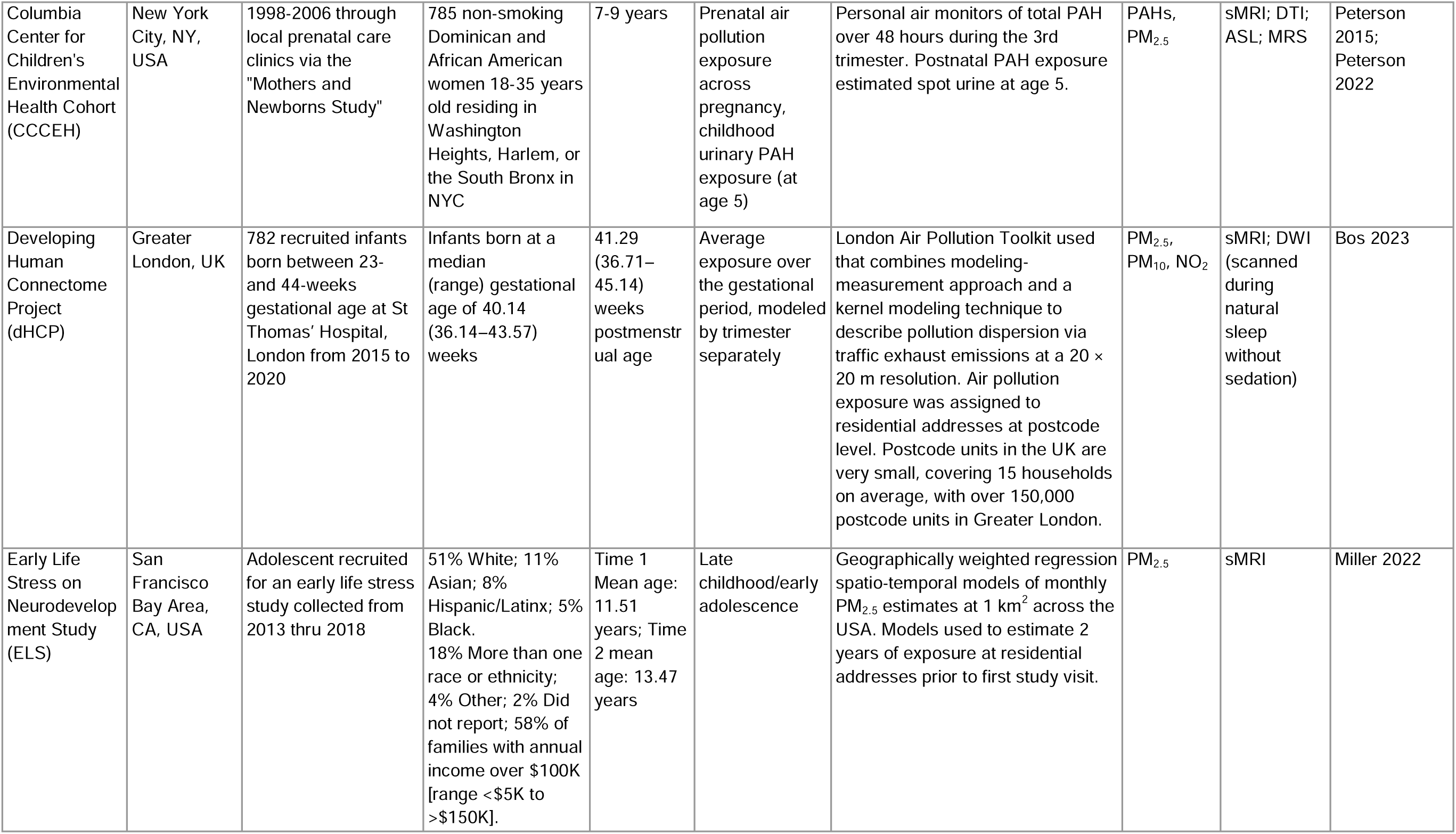
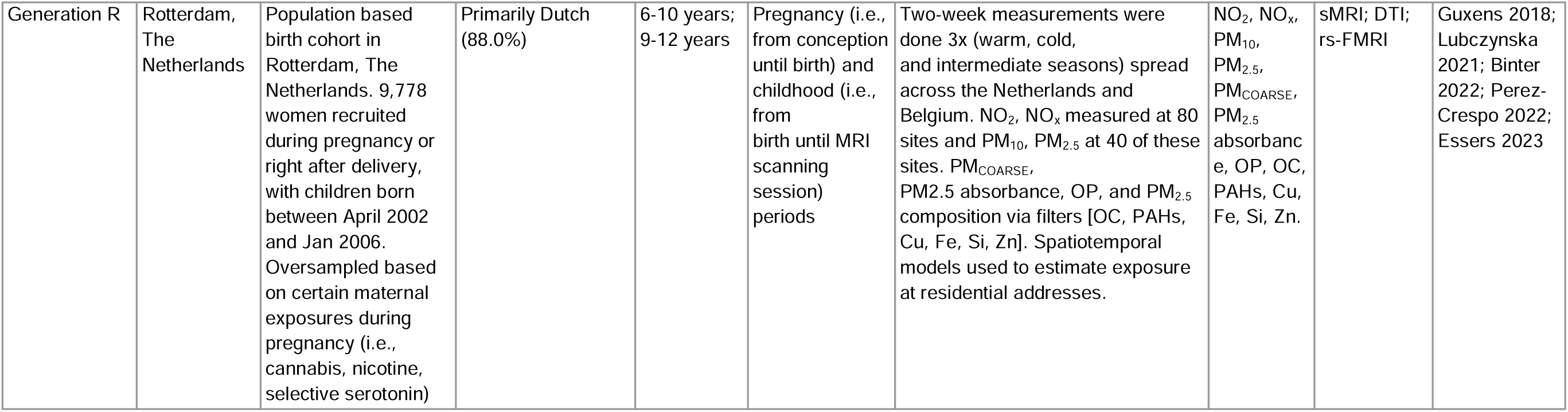
Details of study cohorts included in the current study, participant demographics, exposures examined, and MRI outcomes utilized. Abbreviations: ABCD = Adolescent Brain Cognitive Development Study; BREATHE = Brain Development and Air Pollution Ultrafine Particles in School Children Project; CCAAPS = Cincinnati Childhood Allergy and Air Pollution Study; CCCEH = Columbia Center for Children’s Environmental Health Cohort; dHCP = Developing Human Connectome Project; ELS = Early Life Stress on Neurodevelopment Study; Generation R = Generation R Study. CA = California; OH = Ohio; NY = New York. BC = Black Carbon; Cu = Copper; EC = Elemental Carbon; ECAT = Elemental Carbon Attributable to Traffic; Fe = Iron; NO_2_ = Nitrogen Dioxide; NO_x_ = Nitric Oxide; O_3_ = Ozone; OC = Organic Carbon; OP = Oxidative Potential of PM_2.5_; PAH = Polycyclic Aromatic Hydrocarbon; PM_2.5_ = Fine Particulate Matter; PM_10_ = Coarse Particulate Matter; PM_COARSE_ = Particulate matter between 2.5-10µm; Si = Silicon; UFP = Ultrafine Particulate Matter; Zn = Zinc. ASL = Arterial Spin Labeling; DTI = Diffusion Tensor Imaging; DWI = Diffusion Weighted Imaging; MRI = Magnetic Resonance Imaging; MRS = Magnetic Resonance Spectroscopy; rs-fMRI= Resting State Functional Magnetic Resonance Imaging; RSI = Restriction Spectrum Imaging; sMRI: Structural Magnetic Resonance Imaging.

The **A**dolescent **B**rain **C**ognitive **D**evelopment Study (ABCD Study®) is the largest long-term study of brain development and child health in the United States. This cohort enrolled 9-10 year-old children from 21 study sites from October 2016 through October 2018 across the United States and includes annual study visits, with biennial MRI brain scans. Data from this cohort was used in three cross-sectional (Burnor et al., 2021; Cserbik et al., 2020; Sukumaran et al., 2023) and one longitudinal MRI study (Cotter et al., 2023). All four studies included approximately equal distributions by sex and reported a breakdown of race/ethnicity among participants, with comparable percentages across studies. Most participants identified as Non-Hispanic White, while the fewest participants identified as Asian. The remaining participants identified as Hispanic, Non-Hispanic Black, or multi-racial/other, in descending order of percent represented.

The **Br**ain D**e**velopment and **A**ir Pollu**t**ion Ultrafine Particles in Sc**h**ool Childr**e**n (BREATHE) Project enrolled 7-9 year-old children from 39 schools based in Barcelona, Spain from January 2012 through February 2013. A subset of this cohort underwent neuroimaging at a mean age of 9 years-old, which was reported on in five cross-sectional MRI studies (Alemany et al., 2018; Mortamais et al., 2019, 2017; Pujol et al., 2016b, 2016a). Participants were relatively equally distributed by sex, although neither race/ethnicity nor parental country of birth were reported.

The **C**incinnati **C**hildhood **A**llergy and **A**ir **P**ollution **S**tudy (CCAAPS) is a prospective cohort study that recruited newborns living either 1) <400 meters of an interstate highway or 2) >1 kilometer of an interstate highway in the Greater Cincinnati Metropolitan Region of southwestern Ohio and northern Kentucky from October 2001 through September 2006. Participants were later invited to participate in cross-sectional brain imaging at approximately age 12 years (Beckwith et al., 2020; Brunst et al., 2019). Participants were relatively equally distributed by sex, and the majority identified as White (>70%), with the remaining participants identified as African American or more than one race.

The **C**olumbia **C**enter for **C**hildren’s **E**nvironmental **H**ealth **C**ohort (CCCEH) is a birth cohort study that began in 1998 and enrolled minority mothers from New York City. Children were then invited to participate in cross-sectional brain imaging at either ∼8 years or ∼10 years old (Peterson et al., 2022, 2015). Participants were relatively equally distributed by sex. For both studies, race/ethnicity was reported based on maternal identification, with the majority identifying as Dominican (>60%) and the remainder identifying as African American.

One study by Bos et al. (2023) reported findings from the **D**eveloping **H**uman **C**onnectome **P**roject (dHCP), which aims to map brain connectivity from 22 to 44 weeks post-conception age. This study is conducted in the greater London area of the United Kingdom, with the median age of participants reported as 41.29 weeks postmenstrual age. Infants were born between 2015 and 2020 and underwent a cross-sectional MRI during the neonatal period. There were relatively equal male and female participants, albeit neither race/ethnicity nor parental country of origin was reported for this study.

The **E**arly **L**ife **S**tress on Neurodevelopment (ELS) Study, reported on by Miller et al. (2022), is a longitudinal MRI study based in San Francisco/San Jose Bay Area. The mean age of participants was 11.51 years at baseline and 13.47 years at a two-year follow-up visit. There were a relatively equal number of male and female participants. Participants identified as White, more than one race/ethnicity, Asian, Hispanic, Black, and “Other” in descending order of percent represented.

The Generation R Study is a population-based prospective birth cohort located in Rotterdam, Netherlands, with pregnant women enrolled based on an expected delivery date between April 002 and January 2006. Children from this cohort were later invited for MRI brain imaging between the ages of 6-12 years. This data was utilized for five cross-sectional MRI studies (Binter et al., 2022; Essers et al., 2023; Guxens et al., 2018; Lubczyńska et al., 2021; Pérez-Crespo et al., 2022). Most of these studies had approximately equal distributions of participants by sex, but did not report race/ethnicity, with the exception of Pérez-Crespo et al. (2022). This study reported race/ethnicity based on maternal country of birth, with the majority (>50%) of participants identifying as Dutch. The second identified race/ethnicity was “Nonwestern”, and the last most identified was “Other Western”.

### 3.1.2 Summary of Exposures

The majority of studies estimated exposure levels based on residential address of participants using spatiotemporal modeling (e.g., land use regression, hybrid or ensemble modeling), whereas the CCCEH study obtained measurements from personal air monitors from mothers over a 48-hour period during pregnancy, and the BREATHE Project measured exposure levels via high-volume samplers and passive dosimeters within the courtyards of the participant’s schools (**Figure 3A**).

**Figure 3.**
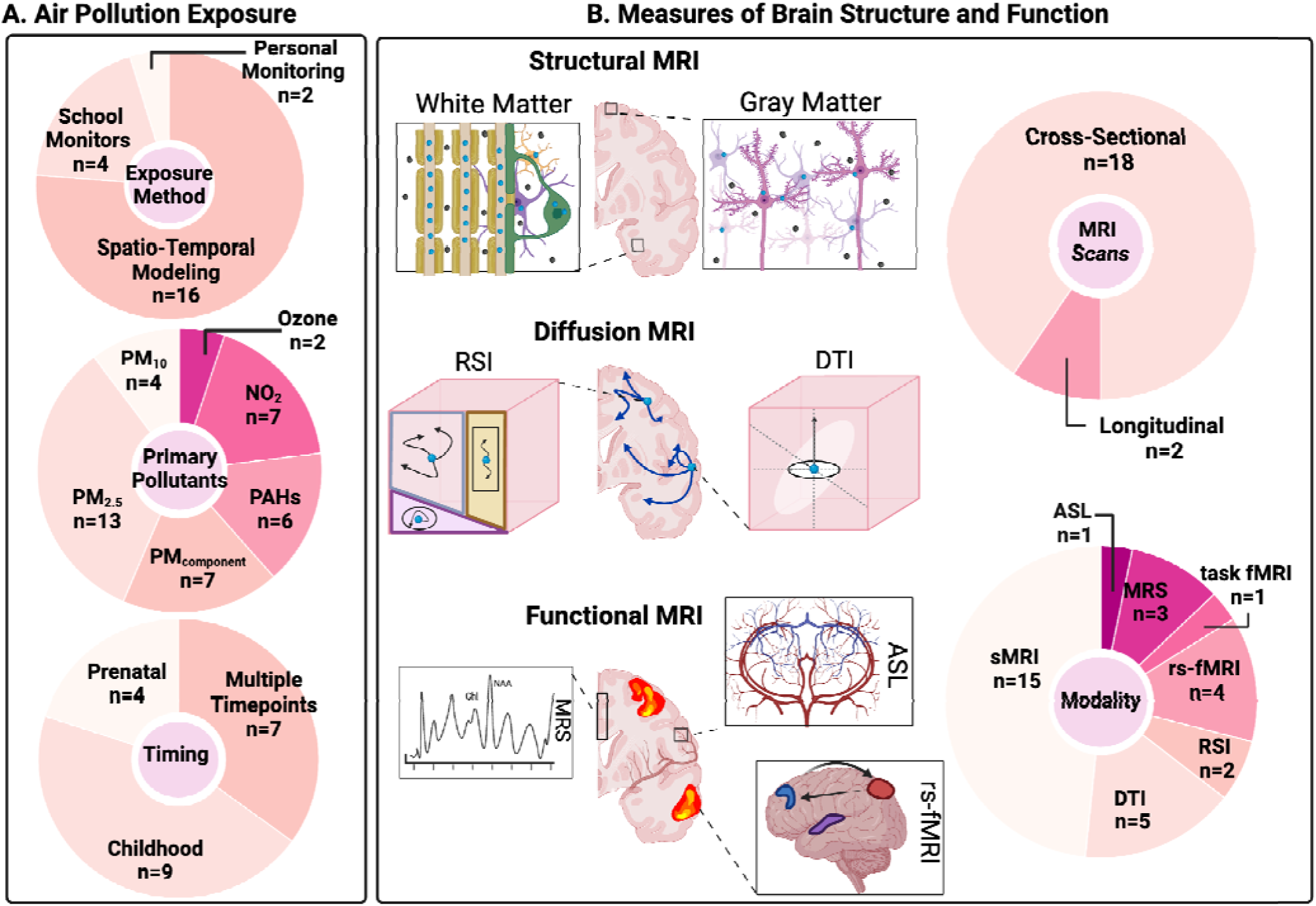
A) Summary of air pollution exposures across studies, including exposure estimation methodology, primary exposure(s), and timing of interest; B) Summary of MRI outcomes for brain structure and function across studies, including MRI modality methodology and study design for MRI outcomes. Abbreviations: NO_2_ = Nitrogen Dioxide; PAH = Polycyclic Aromatic Hydrocarbon; PM_2.5_ = Fine Particulate Matter; PM_10_ = Coarse Particulate Matter. ASL = Arterial Spin Labeling; DTI = Diffusion Tensor Imaging; MRI = Magnetic Resonance Imaging; MRS = Magnetic Resonance Spectroscopy; rs-fMRI= Resting State Functional Magnetic Resonance Imaging; RSI = Restriction Spectrum Imaging; sMRI: Structural Magnetic Resonance Imaging. Created with Biorender.

Using these methods, studies examined a range of pollutants, including two classes of PM (PM_2.5_ and PM_10_), Polycyclic Aromatic Hydrocarbons (PAH), and other gaseous pollutants such as NO_2_, NO_x_, and O_3_ (**Figure 3A**, **Tables 1-2**). Additional PM components were also estimated (n = 7), including elemental carbon (EC; n = 2) or attributable to traffic (ECAT; n = 2), copper (Cu; n = 3), organic carbon (OC; n = 2), iron (Fe; n = 2), potassium (K; n = 1), silicon (Si; n = 2), zinc (Zn; n = 2) and oxidative potential of PM_2.5_ (OP; n = 2).

The timing of exposures examined across the 20 studies included in this review ranged from the prenatal period through early adolescence, with several studies assessing multiple exposure timings in their analyses (n = 7) (**Figure 3A**; **Tables 1-2**). Using the pediatric terminology definitions from the National Institute of Child Health and Development (Williams et al., 2012), across all studies, eight included prenatal exposures and 3 included exposures from birth through toddlerhood. Most studies examined childhood through early adolescence, including early childhood (n = 1), middle childhood (n = 12), and early adolescence (n = 1). Five studies included measures of average childhood exposure; Beckwith et al. (2020), Brunst et al. (2019), Essers et al. (2023), and Lubczyńska et al. (2021) assessed average exposure from birth through age 12 years, and Binter et al. (2022) included an average of exposure from conception until time of MRI (i.e, 9-12 years-old).

### 3.1.3 Summary of Outcomes

Across the 20 included studies, 18 examined brain MRI outcomes cross-sectionally, and two used longitudinal MRI outcomes (Cotter et al., 2023; Miller et al., 2022) (**Figure 3B**). Several structural and functional neuroimaging modalities were leveraged, including sMRI (n = 15), diffusion tensor imaging (DTI; n = 5), restriction spectrum imaging (RSI; n = 2), arterial spin labeling (ASL; n = 1), magnetic resonance spectroscopy (MRS; n = 3), resting-state fMRI (rs-fMRI; n = 4), and task fMRI (n = 1) (**Figure 3B**). Additional details regarding MRI acquisition and imaging preprocessing can be found in Supplemental Table 2.

#### 3.2 Summary of Associations

Across a range of exposure periods, several associations between pollutant exposure(s) and structural and functional brain outcomes were identified (**Table 2**). Across the studies, both single-pollutant models (e.g., assessing a single pollutant in each model), and multi-pollutant models (e.g., several pollutants occur in the same model) (Dominici et al., 2010) were used. The significant findings are summarized below by MRI modality. Given the dynamic changes in brain structure that occur both prenatal and postnatally, we also further discuss these specific findings below based on either prenatal or postnatal exposure periods.

### 3.2.1 Structural MRI (sMRI)

The majority of studies focused on how air pollution exposure relates to the shape and size of gray and white matter using sMRI (**Figure 3B**). Seven studies identified relationships between prenatal air pollution exposure and gray matter volume (n = 4), white matter volume (n = 1), surface area (n = 2), and/or cortical thickness (n = 2); ten studies identified associations between childhood air pollution exposure and gray matter volume (n = 7), white matter volume (n = 1), surface area (n = 1), and/or cortical thickness (n = 2).

#### 3.2.1.1 Prenatal Exposure

One study examined the association between prenatal air pollution exposure and brain structure in infancy (Bos et al., 2023). Higher prenatal PM_10_ concentrations and lower NO_2_ concentrations were associated with larger ventricle size and larger cerebellum, as well as smaller cortical gray matter, amygdala, and hippocampus volumes in infants (median age 41.29 weeks postmenstrual age) from the dHCP study (n = 469; Bos et al., 2023). Several studies, including those pioneering this area of research, have also identified relationships between prenatal exposure and structural brain outcomes in childhood and early adolescence. Prenatal PAH exposure during the third trimester was found to be significantly associated with decreases in white matter surface area, but not gray matter, in children 7-9 years old from the CCCEH cohort (n = 40; Peterson et al., 2015). In an expanded CCCEH sample, prenatal PM_2.5_ and PAH were found to be related to cortical thickness in 6-14 year-olds (n = 332; Peterson et al., 2022), with higher levels associated with thickening in lateral temporal, posterior inferior, and mesial wall surfaces, along with thinning in dorsal posterior surfaces. Both exposures were also associated with smaller white matter surfaces in inferior parietal lobes and cingulate gyrus, and larger white matter surfaces in dorsal convexity, superior frontal gyrus, and posterior inferior brain surface (Peterson et al., 2022). Sex moderated these effects (see details in **section 3.3**). Similarly, higher levels of prenatal PM_2.5_ exposure during the third trimester of pregnancy was associated with a decrease in corpus callosum volumes in 8–12-year-old children from the BREATHE project (n = 186; Mortamais et al., 2019). Although these results remained unchanged in adjusting for current (i.e. childhood) PM_2.5_ exposure, prenatal results did not survive multiple comparison correction (Mortamais et al., 2019).In contrast, another study reported that prenatal PM_2.5_ exposure was associated with gray matter thickness, but not white matter volume, in 6–10-year-olds from the Generation R cohort (n = 783; Guxens et al., 2018). Albeit, in a follow-up study, single pollutant models of exposure in Generation R found that higher prenatal exposure to several air pollutants was associated with smaller corpus callosum and hippocampus volumes, and larger cerebellum, putamen, pallidum, and amygdala at 9-12 years (n = 3,113; Lubczyńska et al., 2021). Moreover, in multi-pollutant models also accounting for postnatal exposure, higher exposure to prenatal oxidative potential (evaluated using electron spin resonance; OP_ESR_) was associated with smaller corpus callosum volume, whereas higher prenatal exposure to PAHs was associated with smaller hippocampus volumes (Lubczyńska et al., 2021).

#### 3.2.1.2 Childhood Exposure

Initial studies of childhood exposure and brain outcomes stemmed from the BREATHE project, which found associations between traffic-related air pollution (TRAP) exposure at school and brain structure in 8-12 year-olds (n = 163; Alemany et al., 2018; n = 242; Mortamais et al., 2017; n = 263; Pujol et al., 2016a). Exposure to several pollutants, including copper, a traffic-related pollution indicator (i.e., derived from elemental carbon and NO_2_), and PAHs, was associated with smaller volumes and less gray matter density in the caudate (Alemany et al., 2018; Mortamais et al., 2017; Pujol et al., 2016a). Following these landmark studies, emerging research continues to suggest childhood exposure may influence various structural brain outcomes depending on timing of the exposure and outcome. Higher levels of one-year of PM_2.5_ exposure during late childhood was associated with both increases and decreases in surface area, cortical thickness, and subcortical volumes at 9-10 years-of-age in participants from the ABCD Study (n = 10,343; Cserbik et al., 2020). Subcortical volumetric findings included higher exposure to be linked to larger right thalamus, right pallidum, and left accumbens volumes, but smaller left putamen and left pallidum volumes. Using lifetime estimates of exposure, several exposures and structural brain differences at 9-12 years-old have also been noted in the Generation R cohort. In single pollutant analyses, higher exposure to some pollutants from birth to time of MRI was associated with larger nucleus accumbens and smaller corpus callosum and hippocampal volumes (n = 3,113; Lubczyńska et al., 2021). In multi-pollutant models accounting for prenatal exposure, higher postnatal exposure to organic carbon (OC) was associated with a smaller corpus callosum, whereas higher exposure to oxidative potential (evaluated using dithiothreitol; OP_DTT_) was associated with a smaller hippocampus (Lubczyńska et al., 2021). Using distributed lag modeling to identify sensitive periods of exposure, Binter and colleagues found that greater PM_2.5_ exposure in infancy (4 months to 1.8 years) was associated with larger putamen volume in the Generation R cohort (n = 3,515; Binter et al., 2022). A CCAPS cohort study also suggested the importance of lifetime exposure on gray matter morphology, as cortical thickness at age 12 was reduced in those exposed to higher versus lower ECAT exposure from birth to age 12 years-old (n = 135; Beckwith et al., 2020) regions showing reductions included bilateral paracentral lobule, postcentral gyrus, superior frontal gyrus, superior parietal lobule, cerebellum; left precentral and left precuneus; as well as right posterior cingulate and right parietal cortex (Beckwith et al., 2020). Lastly, the ELS study found 2 years of PM_2.5_ exposure was associated to longitudinal changes in structural MRI over a 2-year period from ages 9-13 years-old, including gray matter expansion and white matter volumes in a region- and hemispheric-specific fashion as measured by voxel based morphometry (n = 115; Miller et al., 2022). Independent of early life stress (see details in **section 3.3**), higher exposure was positively associated with gray matter within the left precentral gyrus and cerebellum, bilateral medial orbitofrontal cortex, as well as white matter estimates in the left caudate, left corpus callosum, left cingulum, bilateral inferior fronto-occipital fasciculus, and right inferior frontal and temporal gyrus in 9-13 year-olds. In addition, PM_2.5_ was also negatively associated with gray matter in the left insula, left cerebellum, right cingulate gyrus, and caudate, as well as white matter estimates in the left inferior temporal gyrus, angular gyrus, posterior thalamic radiation, middle frontal gyrus, hippocampal cingulum, and right post central gyrus.

#### 3.2.1.3 Summary of sMRI Findings

All seven prenatal studies identified relationships between exposure to PM or its components (e.g., PAH) and brain morphology, suggesting that the prenatal period is vulnerable to the neurotoxic effects of PM. The ten childhood and adolescent studies also highlight how continuing postnatal brain development may be impacted by air pollution, including longitudinal changes in gray and white matter development during early adolescence. Albeit, specific pollutants, brain regions, and directionality of associations were largely mixed across studies (see **Figure 4** for summary of commonly identified brain regions), suggesting region-specific associations between air pollution and structural brain outcomes likely depend on the specific timing, duration, and pollutant profile of the exposure.

**Figure 4.**
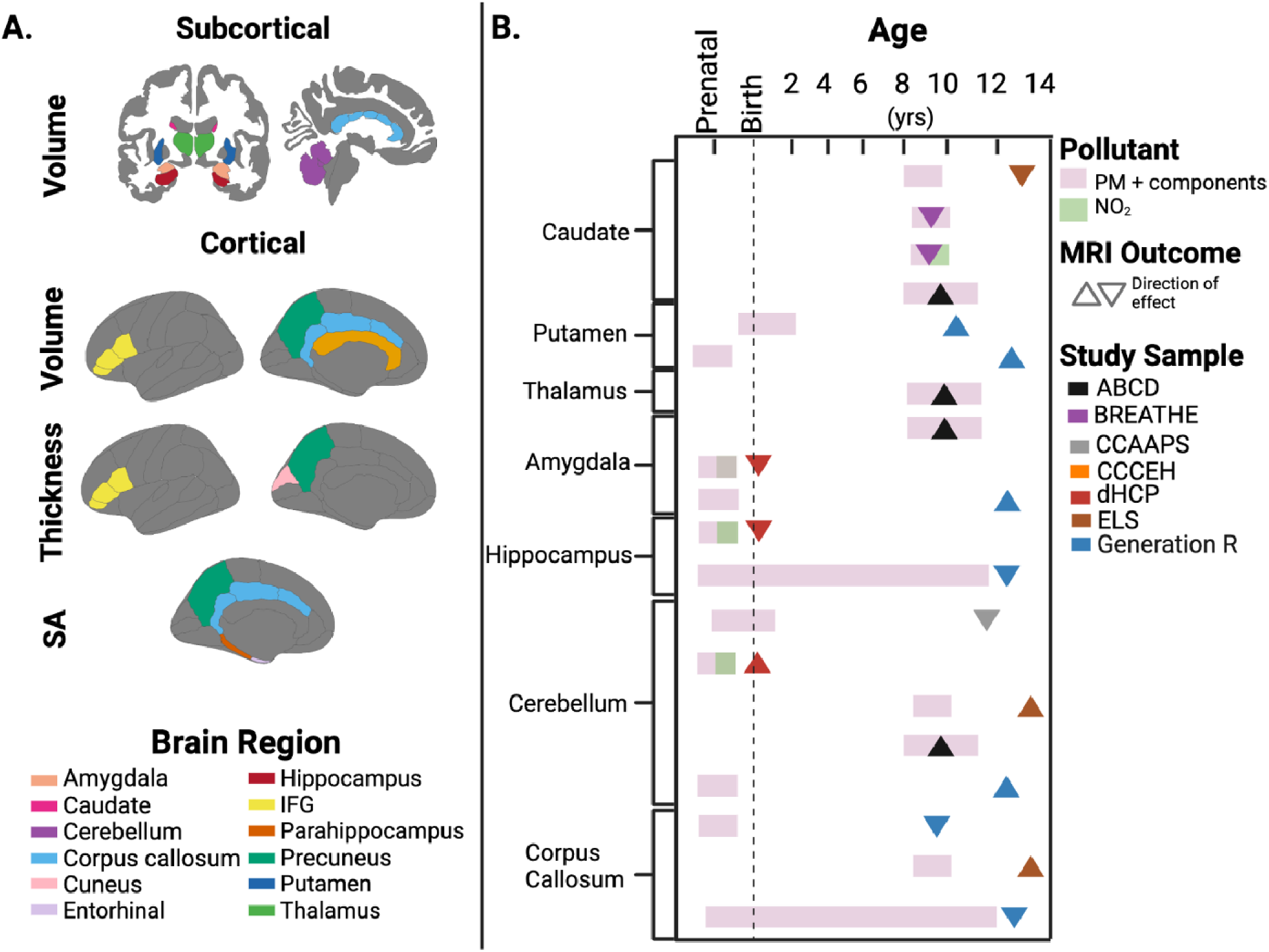
A) Structural Magnetic Resonance Imaging (MRI) brain regions commonly identified across multiple studies. B) Illustration of variable timing of exposure and volumetric outcomes across select studies including in review. Summary of MRI outcomes for brain structure and function across studies, including MRI modality methodology and study design for MRI outcomes. Abbreviations: IFG = inferior frontal gyrus; SA = Surface Area. PM = particulate matter; NO_2_ = nitrogen dioxide. ABCD = Adolescent Brain Cognitive Development Study; BREATHE = B**r**ain Development and Air Pollution Ultrafine Particles in School Children Project; CCAAPS = Cincinnati Childhood Allergy and Air Pollution Study; CCCEH = Columbia Center for Children’s Environmental Health Cohort; dHCP = Developing Human Connectome Project; ELS = Early Life Stress on Neurodevelopment Study; Generation R = Generation R Study. Created with Biorender.

### 3.2.2 Diffusion MRI (dMRI)

DMRI measures the diffusion of water molecules in biological tissues (Lebel and Deoni, 2018). Depending on acquisition sequence complexity and water diffusion modeling, various diffusion metrics allow for measurement of tissue properties, thought to reflect various biological properties in white matter and gray matter. **Figure 5** highlights the 6 studies that have examined air pollution exposure and tissue microstructure using dMRI. Of these studies, 4 used diffusion tensor imaging (DTI), which uses three primary directions and magnitudes of the diffusion coefficient to map tissue structure (O’Donnell and Westin, 2011), and 2 used restriction spectrum imaging (RSI), which is an advanced multi-compartment modeling technique that can differentiate between different types of intra- and extracellular diffusion (Brunsing et al., 2017). DTI outcomes of interest include extracellular diffusion along a single direction, known as fractional anisotropy (FA) and an overall degree of diffusion in any direction, known as either mean diffusivity (MD) or the apparent diffusion coefficient (ADC). Higher FA and lower MD/ADC may represent increased myelination, axonal packing, and/or fiber coherence; MD can be used to assess gray matter microstructure, with an increase in MD potentially indicating a breakdown of tissue barriers, and thus reduced gray matter volume (Feldman et al., 2010; Weston et al., 2015). RSI outcomes of interest within the air pollution literature so far include restricted directional (RND) and isotropic (RNI) intracellular diffusion. RND represents intracellular diffusion in a single direction while RNI represents intracellular diffusion in any direction. Higher RND may indicate increased axonal but caliber, density, and/or myelination in white matter and greater neurite organization and/or density in gray matter, while higher RNI potentially represents larger or increased quantities of cell bodies in both white and gray matter (Palmer et al., 2022; White et al., 2013). Using these approaches, only one study identified significant relationships between prenatal exposure and dMRI metrics (Peterson et al., 2022), whereas four studies found associations between childhood exposure and dMRI outcomes (Binter et al., 2022; Burnor et al., 2021; Pujol et al., 2016a; Sukumaran et al., 2023).

**Figure 5.**
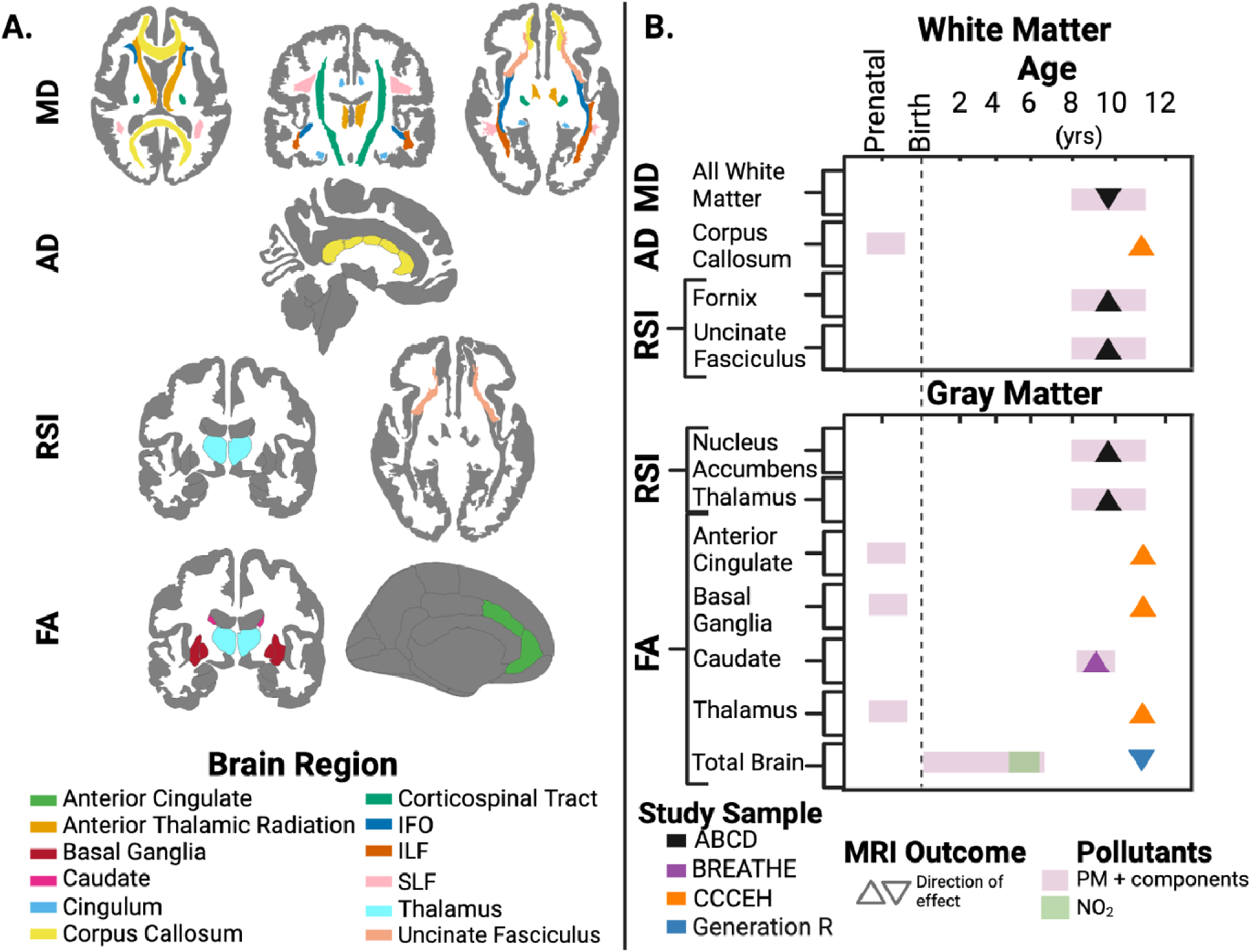
A) Diffusion-weighted MRI brain regions commonly identified across multiple studies. B) Illustration of variable timing of exposure and diffusion-weighted imaging outcomes across all five included studies. Summary of MRI outcomes for brain structure and function across studies, including MRI modality methodology and study design for MRI outcomes. Abbreviations: AD = axial diffusivity; FA = fractional anisotropy; IFO = inferior longitudinal occipital; ILF = inferior longitudinal fasciculus; MD = mean diffusivity; RSI = restricted spectrum imaging; SLF = superior longitudinal fasciculus. PM = particulate matter; NO_2_ = nitrogen dioxide. ABCD = Adolescent Brain Cognitive Development Study; BREATHE = B**r**ain Development and Air Pollution Ultrafine Particles in School Children Project; CCCEH = Columbia Center for Children’s Environmental Health Cohort; Generation R = Generation R Study. Created with Biorender.

#### 3.2.2.1 Prenatal Exposure

In the CCCEH cohort, prenatal PM_2.5_ during the 3rd trimester was associated with a greater ADC in white matter at 6-14 years old, including posterior white matter tracts, anterior corpus callosum, and anterior corona radiata, whereas PAH was associated with a lower ADC in the interior capsule, driven by axial diffusivity (AD) (n = 332; Peterson et al., 2022). In gray matter tissue, prenatal exposure to both PM_2.5_ and PAH were found to be independently associated with higher FA in the basal ganglia, thalamus, and anterior cingulate gyrus (Peterson et al., 2022).

#### 3.2.2.2. Childhood Exposure

Pujol et al. (2016a) identified a positive relationship between one year of copper exposure at school and FA in the caudate nucleus and the adjacent white matter tract in 8-12 year-olds from the BREATHE cohort (n = 263). Since then, two large cohort studies have found associations between postnatal PM_2.5_ exposure and white matter microstructure. One year of childhood PM_2.5_ exposure was negatively associated with white matter MD, but positively, albeit non-linearly, associated with RNI in left cingulum, left superior longitudinal fasciculus, and bilaterally in fornix and uncinate fasciculus in 9-10 year-olds from the ABCD Study (n = 7,602; Burnor et al., 2021). A second study in the Generation R cohort found various patterns of exposure at different periods of development related to lower global brain FA at 9-12 years-old, including higher NO_2_ during early childhood (i.e., 3.6-4.8 years), higher prenatal and early childhood PM_2.5_ (i.e., conception to 4 years), and higher PM_2.5_ absorbance early in life (i.e., estimate of elemental/black carbon from birth to 5 years) (n = 3,515; Binter et al., 2022). Lastly, in terms of gray matter microstructure, positive associations were found between childhood annual PM_2.5_ exposure and RNI of bilateral nucleus accumbens, right thalamus, and the brainstem in 9-10 year-olds from the ABCD Study, while also accounting for co-exposure to NO_2_ and O_3_ using a multivariate analytic approach (n = 8,796; Sukumaran et al., 2023).

#### 3.2.1.3 Summary of dMRI Findings

Patterns and directionality of associations across the one prenatal and four childhood air pollution exposure studies were again mixed, potentially due to study-to-study differences in specific pollutants, timing of exposure, and methodological dMRI differences utilized across studies (e.g., multi-shell DTI vs. single shell-DTI; **Supplemental Table 2**). However, all five studies identified significant associations between PM_2.5_ and tissue microstructure, further suggesting that microarchitecture of the developing brain might be particularly susceptible to the effects of PM_2.5_ and its components.

### 3.2.3 Functional MRI (fMRI)

FMRI estimates brain activity by measuring changes in the blood-oxygen-level-dependent (BOLD) signal over time (Hillman, 2014). Resting-state fMRI (rs-fMRI) studies examine BOLD signal correlations between brain regions while a participant is at rest, whereas task-based fMRI studies look at changes in the BOLD signal when a participant engages in a behavioral task (e.g., listening to a song or viewing a series of pictures) as compared to a control task (i.e., simplified sensory or motor task; fixation cross) (Shah et al., 2010). With regards to the functional implications of air pollution exposure on the brain, four studies identified associations between pollutants and functional connectivity (FC; Cotter et al., 2023; Pérez-Crespo et al., 2022; Pujol et al., 2016b, 2016a), while one of these studies also identified a relationship between air pollutant exposure and brain activation to sensory stimuli (Pujol et al., 2016b).

#### 3.2.3.1 Prenatal Exposure

One study from the Generation R cohort assessed relationships between prenatal exposure to TRAP and road traffic noise and FC at 9-12 years-of-age; however, no significant associations were identified when examining the prenatal exposure period (n = 2,197; Pérez-Crespo et al., 2022).

#### 3.2.3.2 Childhood Exposure

The first fMRI studies were conducted in 7-9 year-old participants from the BREATHE Cohort, which found exposure to copper and a TRAP indicator (i.e., derived from elemental carbon and NO_2_) at school was associated with altered functional brain activation to sensory stimuli (Pujol et al., 2016b), as well as resting-state FC networks (n = 263; Pujol et al., 2016a, 2016b). Since those initial studies, two additional rs-fMRI studies have been published. In a cross-sectional study from Generation R, early life exposure to NO_2_ (i.e., birth to 3 years) was related to inter-network FC between the superior parietal cortex and dorsolateral prefrontal cortex, and between auditory association cortex and inferior frontal cortex in 9–12 year-olds (n = 2,197; Pérez-Crespo et al., 2022).Early childhood exposure to NO_x_ (i.e., ages 3 to 6 years) was also associated with inter-network, intra-hemispheric FC between inferior frontal cortex and medial temporal cortex and between inferior and frontal opercular cortex and medial temporal cortex. Finally, early life PM_2.5_ absorbance (i.e., EC exposure from birth to 3 years) was also associated with many mostly inter-network FC between brain regions belonging to task positive and task negative networks (Pérez-Crespo et al., 2022). More recently, one longitudinal study examined associations between one-year childhood exposure to criteria pollutants, NO_2_, O_3_, and PM_2.5_ and resting-state changes over a 2-year follow-up period from ages 9-13 years in the ABCD Study cohort (n = 9,497; Cotter et al., 2023). In multi-pollutant models, associations between PM_2.5_ exposure and inter-network FC development over time were noted for salience to default mode, and frontoparietal to default mode networks. Higher PM_2.5_ exposure was also associated with differences in amygdala and hippocampal FC to frontoparietal, default mode, and salience networks longitudinally from 9-13 years-old. Multi-pollutant models also suggested higher NO_2_ and O_3_ exposure were associated with additional within-network integration and between-network segregation over the two-year follow up period (Cotter et al., 2023).

#### 3.2.3.3 Summary of fMRI Findings

Overall, results from these four studies suggest that childhood air pollution exposure may be associated with altered patterns of sensorimotor brain activity and large-scale functional network organization, including how these patterns change longitudinally with development during early adolescence.

### 3.2.4 MR Spectroscopy

Three studies have utilized magnetic resonance Spectroscopy (MRS), a method used for *in-vivo* imaging of biochemical compounds and measurement of brain metabolism (Zhu and Barker, 2011). All studies acquired MRS, with one study using a whole-brain approach above the anterior to posterior commissure (Peterson et al., 2022), one focusing on the frontal white matter (Pujol et al., 2016b), and the third focusing specifically on the cingulate (Brunst et al., 2019).

#### 3.2.4.1 Prenatal Exposure

In the CCCEH cohort, prenatal exposure to both PM_2.5_ and PAH was associated with higher N-acetylaspartate (NAA) concentrations in the dorsal anterior cingulate cortex (dACC) and higher choline concentrations in dACC and midcingulate cortex at 6-14 years (n = 332; Peterson et al., 2022). Prenatal PAH exposure alone was also associated with higher NAA and choline concentrations in pregenual ACC, insula, centrum semiovale, and corona radiata (Peterson et al., 2022).

#### 3.2.4.2 Childhood Exposure

In 12 year-olds from CCAAPS, higher myo-inositol within the ACC were seen in those exposed to one-year of high versus low ECAT exposure (n = 145, CCAAPS; Brunst et al., 2019). Pujol and colleagues (2016b) found no significant associations between childhood exposure and brain metabolism.

#### 3.2.3.3 Summary of MRS Findings

These initial studies suggest that brain metabolism within portions of the cingulate cortex might be particularly vulnerable to air pollution exposure during both the prenatal and postnatal periods.

### 3.2.5 Arterial Spin Labeling (ASL)

ASL is an imaging technique that uses magnetically labeled water protons that are found in arterial blood to assess the rate of blood flow to brain tissues (Iutaka et al., 2023). Only a single study has examined air pollution and cerebral blood flow (CBF), as measured by ASL.

#### 3.2.5.1 Prenatal Exposure

In the CCCEH cohort, higher levels of prenatal exposure to both PM_2.5_ and PAH were related to lower relative CBF (rCBF) in frontal gray matter at 6-14 years-old (n = 332; Peterson et al., 2022). In addition, exposure to PM_2.5_ was negatively associated with rCBF in the lingual and fusiform gyri, hippocampus, and dorsal parietal cortex. Finally, PAH exposure was negatively associated with rCBF in the putamen, thalamus, and midcingulate cortex, and white matter of internal and external capsules, corpus callosum genu, and anterior corona radiata. Again, sex moderated these effects (see **section 3.3**).

#### 3.2.5.2 Childhood Exposure

No studies have examined associations between childhood air pollution exposure and ASL outcomes.

#### 3.2.3.3 Summary of ASL Findings

The initial ASL study suggests prenatal exposure may relate to hypoperfusion in various brain regions during childhood and adolescence.

##### 3.3 Modifying effects of genetics, early life stress, and sex

A few studies have investigated whether air pollution effects on sMRI and dMRI are moderated by various genetic, biological, and environmental factors. In one of the first studies, Alemany and colleagues investigated whether associations between air pollution exposure and brain volumes were moderated by apolipoprotein E (APOE) ε4 status–a genetic risk factor for Alzheimer’s Disease (AD) in the BREATHE cohort (n = 163; Alemany et al., 2018). Results suggested that the negative associations between annual NO_2_ and PAH exposure and caudate volumes were only seen in 8-12 year-olds with the APOE ε4 allele and not in non-carriers (Alemany et al., 2018). A more recent study from Generation R investigated whether the relationships between prenatal air pollution exposure and brain volumes were moderated by multiple genetic risk factors for AD (i.e., APOE ε4 status and AD polygenic risk score (PRS)) in 9-12 year-old children (n = 1,186; Essers et al., 2023). In single pollutant analyses, prenatal and childhood air pollution exposure to several pollutants were differentially associated with various white and gray matter volumes in APOE ε4 carriers compared to non-carriers and for each incremental increase in PRS for AD. In multi-pollutant analyses, only PM_COARSE_ (i.e., PM between 2.5 and 10 μm) exposure during pregnancy and PAH exposure during childhood remained differentially associated with larger cerebral white matter volume in APOE ε4 carriers compared to non-carriers, whereas PM_COARSE_ was only differentially associated with larger cortical gray matter volume per incremental PRS increase for AD (Essers et al., 2023). In addition to genetics, air pollution effects on gray and white matter morphometry over a two-year follow-up period were shown to be modified by early life stress in the ELS Study (n = 115; Miller et al., 2022). In particular, the majority of PM_2.5_ effects were diminished in those who experience early life adversity, potentially suggesting that early life stress may reduce one’s sensitivity to other environmental exposures (Miller et al., 2022). Lastly, several studies have examined potential sex differences in air pollution effects on brain outcomes. In the CCCEH cohort, sex differences were seen in the associations between prenatal exposure and cortical thickness and white matter, with PM_2.5_ associations found to be stronger in boys (Peterson et al., 2022), while PAH associations were stronger in girls (Peterson et al., 2022, 2015). Furthermore, associations between prenatal PM_2.5_ exposure and rCBF in lingual and fusiform gyri, hippocampus, and dorsal parietal cortex were significantly stronger in girls (Peterson et al., 2022). Alternatively, many studies have reported no evidence of sex specific effects for sMRI (Beckwith et al., 2020; Bos et al., 2023; Cserbik et al., 2020; Mortamais et al., 2017, 2019), dMRI (Binter et al., 2022; Burnor et al., 2021) and rs-fMRI (Pérez-Crespo et al., 2022).

##### 3.4 Summary of Bias

Risk of bias summarized across the six domains are reported in **Figure 6**. Each of the domains per study can be found in **Supplemental Figure 2**. Most studies had four or more domains considered to be at ‘low’ risk of bias, with outcome measurement and selective reporting representing the highest proportion of low risk of bias. However, all studies had at least one domain reviewed as ‘moderate’ risk, with missing data representing the highest proportion of moderate risk of bias. Two out of the 20 studies were deemed to be at high risk of confounding bias by not adjusting for all critical confounders identified by the WHO (i.e. SES, age, sex) (Brunst et al., 2019; Peterson et al., 2022). Of the remaining 18 studies, seven were deemed to be at ‘moderate’ risk, due to adjusting for all critical confounders, but not other potential confounders, such as race and/or ethnicity (Alemany et al., 2018; Mortamais et al., 2019, 2017; Pujol et al., 2016b, 2016a) or performing imputation for a large number of missing values for confounding variables (Binter et al., 2022; Pérez-Crespo et al., 2022). The remaining 11 studies were at ‘low’ risk, as all critical and potential confounding variables were identified or included evidence of minimal risk of severe confounding. Eight of the 20 studies were at ‘moderate’ risk of selection bias due to unequal opportunity for participants at all exposure levels to be included in the study (Alemany et al., 2018; Beckwith et al., 2020; Brunst et al., 2019; Essers et al., 2023; Mortamais et al., 2017, 2019; Pujol et al., 2016b, 2016a). Brunst et al. (2019) and Beckwith et al. (2020) used data from the CCAAPS study, which only included children who resided either <400 meters (high TRAP) or >1500 meters (low TRAP) from a major roadway. Several studies used data from the BREATHE study, which specifically recruited low and high NO_2_ schools (Alemany et al., 2018; Mortamais et al., 2019, 2017; Pujol et al., 2016b, 2016a). Essers et al. (2023) only included mothers whose children had available genotypic data and were of European ancestry due to assessment of the *APOE* genotype. The remaining 12 studies were at ‘low’ risk of selection bias due to equal opportunity for participants of all levels of exposure to be studied. All 20 studies were deemed to be at ‘low’ risk of exposure assessment bias due to changing spatial exposure contrasts due to unchanging spatial exposure contrasts throughout the study or using time varying exposure to adjust for changes. One study used a lower level of granularity–postal codes– to estimate air pollution exposure, however the authors state the postal codes encompass less than 15 households (Bos et al., 2023). In articles from the ABCD Study cohort, there is moderate exposure misalignment, resulting from air pollution estimates being calculated for 2016 while some participants completed their MRI visit in 2017-2018 (Burnor et al., 2021; Cotter et al., 2023; Cserbik et al., 2020; Sukumaran et al., 2023). All 20 studies were deemed to be at a ‘low’ risk of outcome measurement bias and selective reporting bias. However, 19 of 20 studies were at ‘moderate’ risk of bias for missing data, since there was >10% loss of participants. However, rationale for this attrition—exclusion, imaging issues, or missing air pollution exposure data—was properly explained in each study. The remaining study was deemed to be at low risk of bias due to having <10% loss of participants related to outcome or exposure data (Binter et al., 2022).

**Figure 6.**
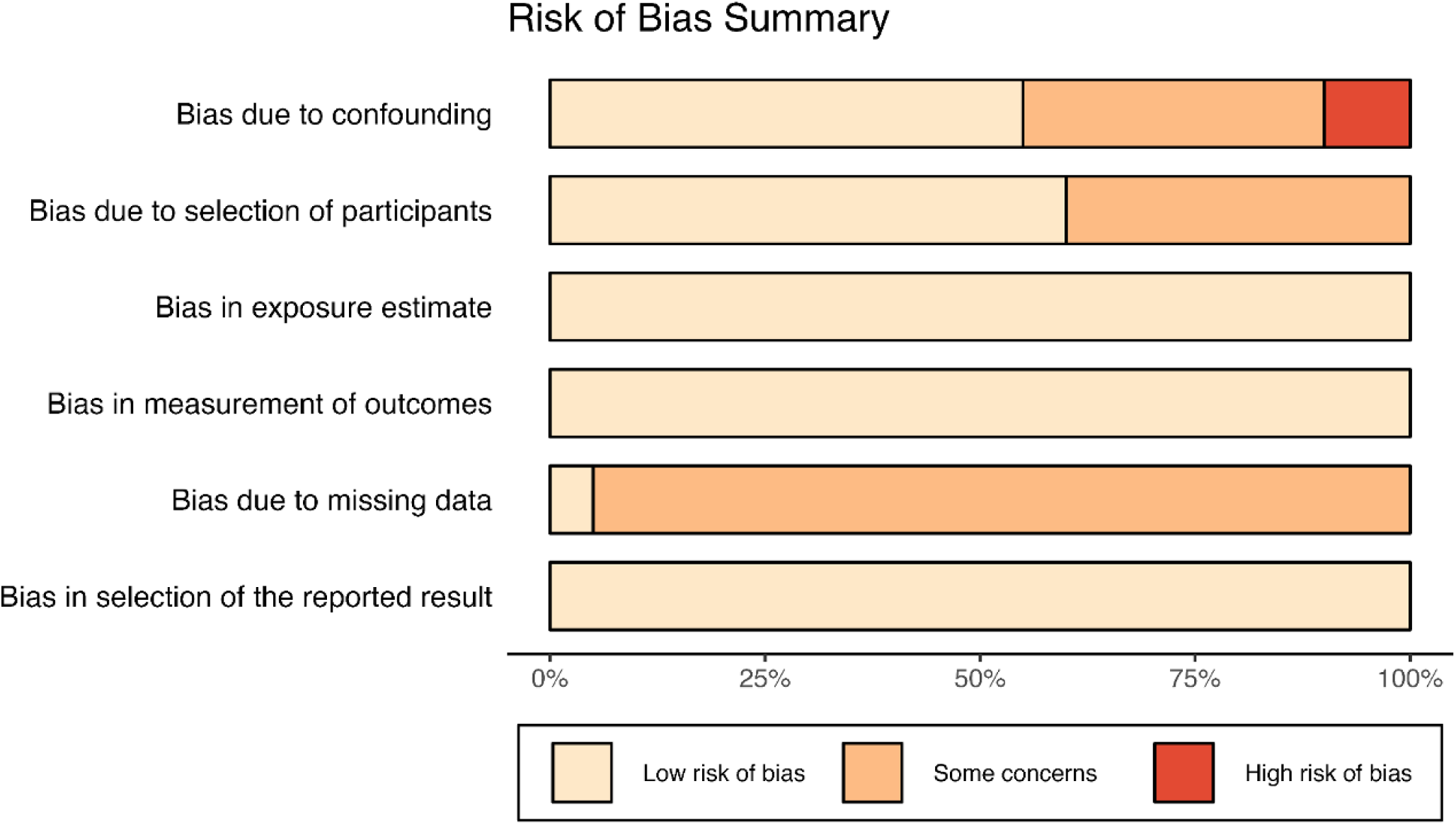
Risk of bias summary. Risk of bias was determined using the WHO Global Air Quality Guidelines. In the figure above, tan indicates low risk of bias, orange indicates some concerns regarding potential bias, and red indicates high risk of bias. Most studies included in this review were found to have low to moderate risk of bias, mostly due to missing data, while a select few demonstrate high risk of bias due to confounding factors.

## 4. Discussion

The current review included 20 MRI studies that explored the relationships between outdoor air pollution exposure and childhood and adolescent brain structure and function. Building on our initial systematic review of the six previously identified studies (Herting et al., 2019), 14 additional studies have been published, including five with lifetime exposure estimates and two with longitudinal MRI outcomes. This considerable growth in the field has been accompanied by substantially increasing sample sizes: the largest includes data from 10,343 participants, and seven additional studies each include well over 1,000. This is important as larger, more diverse samples in neuroimaging studies are critical for statistical power, generalizability, and reproducibility of findings (Kopal et al., 2023; Neuroscience, 2020). However, the 20 studies included are based on data from only seven study cohorts, all from Western countries. Risk of bias across studies was relatively low, apart from two categories: selection bias and bias due to missing data. This highlights the need for additional studies to minimize sampling bias and ensure greater global generalizability.

The cerebellum, corpus callosum, and caudate are among regions most identified by sMRI in the literature, albeit displaying mixed patterns of associations between exposure and brain outcome (**Figure 4B**). While specific brain regions were inconsistent, associations between air pollution and dMRI outcomes was detected in all five studies (**Figure 5B**). Few studies have implemented ASL or MRS; albeit initial studies suggest air pollution may influence both cerebral blood flow and metabolite levels during development. Across modalities, the pollutant most commonly associated with brain outcomes was PM_2.5,_ or components directly derived from it, suggesting that it may be particularly harmful for the developing brain. However, the range of exposure windows, pollutants, and potentially affected brain outcomes makes it difficult to draw strong conclusions from so few studies. Discrepant findings may reflect differential susceptibility to neurotoxic effects of air pollutants across the brain or distinct neurodevelopmental trajectories coinciding with critical periods of vulnerability to environmental insult (Rice and Barone, 2000). Much more research is needed to determine which brain regions, pollutants, and exposure timings exhibit the greatest neurotoxicant effects during development. Below we outline a few key considerations for further study in this emerging area of research.

### 4.1 Timing of Exposure and Brain Outcome

Studies reviewed here investigated a broad range of exposure periods, spanning pregnancy through early adolescence. Differences in the timing of exposure assessments and brain imaging across studies make it difficult to synthesize the current literature. Moreover, while one study scanned participants at 10 months-old, the next youngest participant in any other study was 6 years-old at the time of MRI, whereas the oldest ages of participants were 12 years-old. Hence, there are several years of early development and mid-to-late adolescence during which there are no published studies. In addition, most MRI studies primarily assessed cortical morphology via sMRI, and questions remain about the implications of exposure for microstructural and functional brain development, as well as cerebral blood flow and brain metabolism.

Developmental trajectories of brain structure, function, and blood flow suggest neurotoxicants might affect the brain differently depending on the developmental timing of exposure and the period in which the brain is assessed (Herting et al., 2024). White matter, cortical gray matter, subcortical gray matter, and ventricle volumes, as well as cortical thickness and surface area all follow distinct patterns of growth from birth through early adulthood. For instance, while cortical gray matter volume peaks in mid-childhood, white matter volume continues to increase until young adulthood (Bethlehem et al., 2022). During childhood, cerebral blood flow increases linearly (Paniukov et al., 2020). Functional neurodevelopment is more complex across development, with both integration and segregation of brain networks taking place (Fair et al., 2007). In early life, there is an increase in network segregation followed by a period of integration that persists through adolescence, as network refinement takes place (Edde et al., 2021); albeit these functional studies are largely limited by inferences based on cross-sectional data. Regardless, given dynamic patterns in brain maturation, pollutant(s) are likely to have differential neurotoxic effects depending on the timing of the exposure. This may help to explain how two studies that estimated PM_2.5_ exposure and basal ganglia volumes found opposing effects. Specifically, greater early life exposure (i.e., 4 months to 1.8 years) to PM_2.5_ was linked to greater putamen volume in 9-12 year-olds (Binter et al., 2022), but higher childhood exposure to PM_2.5_ exposure was linked to smaller left putamen volumes at 9-10 years (Cserbik et al., 2020). In the context of previous research demonstrating that basal ganglia volumes peak around 5 years-of-age (Alex et al., 2023), one explanation for the observed opposing associations could be that the basal ganglia may be particularly sensitive to the inflammatory effects of PM_2.5_ during early development, but that when exposure occurs during late childhood/early adolescence, the effects of pruning exert more influence. However, this speculation requires additional research.

Given that timing likely matters for both exposures and brain outcomes, air pollution exposure over various periods of development and repeated brain measurements are essential to more fully characterize air pollution’s neurodevelopmental effects. To this end, only five studies have investigated lifetime air pollution exposure, which is necessary for elucidating impacts of chronic and cumulative pollutant exposure on brain structure and function during late childhood and early adolescence. Across these studies, however, there is no clear consensus on the ages most susceptible to exposure-related neurotoxicity. Yet, they further support the notion that different vulnerable periods may exist for different neurodevelopmental processes. For instance, in the Generation R cohort, differences in global white matter microstructure at ages 9-12 years were linked to PM_2.5_ absorbance from birth to 5 years-old (Binter et al., 2022), whereas differences in inter-network functional connectivity were linked to either exposure estimates from birth to age 3 (NO_2_, PM absorbance) or age 3 to 6 (NO_x_), depending on pollutant (Pérez-Crespo et al., 2022). Alternatively, two studies included longitudinal MRI data, though they only examined how relatively recent periods of air pollution exposure (i.e., 1-2 years prior to first MRI scan) relate to patterns of change in structural morphometry (Miller et al., 2022) and resting-state functional connectivity (Cotter et al., 2023) during early adolescence (∼9-13 years-old). Thus, additional research is necessary that implements a longitudinal approach to studying both exposure and outcome to clarify the developmental impact of air pollution from birth through the end of adolescence.

### 4.2 Sample Demographics and Individual Differences in Susceptibility

The 20 studies reviewed use data from seven unique cohorts. Given many of these are large, longitudinal cohort studies, the wealth of data and insight gained from them are a valuable contribution to this emerging research area, with greater statistical power and participant diversity than is common elsewhere in the pediatric neuroimaging literature. However, there may be a false sense of independence of papers utilizing the same cohort, leading to overestimation of findings that may not replicate in data from other cohorts and thus, may not generalize to the broader population (Mroczek et al., 2022). Furthermore, while these large studies include diverse populations from different geographic regions, all 4 countries represented are Western countries with relatively low pollution levels (PM_2.5_; 5.1 - 10 ug/m^3^) by the World Health Organization (**Figure 2B**). Notably, this review did not identify any studies based in India or China, though these countries are the first and second most populous in the world and have high levels of air pollution (Health Effects Institute, 2024). Because there has not been any research conducted in Africa, Asia, or South America, or in any countries with high levels of outdoor air pollution, the generalizability of findings is limited.

The exclusion of groups most highly affected by air pollution is not only a global issue, but also a major issue in the United States. While economically and socially marginalized communities are disproportionately exposed to neurotoxicants such as air pollution, these groups have been historically excluded from research (Payne-Sturges et al., 2023). Even within counties, different zip codes may have unequal levels of air pollution exposure, often stratified by SES and race (Knobel et al., 2023). Future studies would benefit from assessing exposure levels on a smaller scale, specifically from marginalized communities that are at risk for higher exposure levels and have been historically underrepresented in research. Data gleaned from these analyses could be used to further stratify cohorts, more clearly map the relationship between pollution exposure and adolescent brain structure and function and provide a more generalizable picture of exposure-related consequences in the developing brain.

Individual susceptibility to deleterious effects of outdoor air pollution must also be considered. Three studies assessed environmental or genetic factors moderating exposure-related brain differences, with one study finding air pollution effects on the brain were reduced in individuals with higher early life stress (Miller et al., 2022). This may be due to shared biological pathways–potentially a neuroimmunological link–between environmental and psychological stressors (for review see Olvera Alvarez et al., 2018). Indeed, research has shown that air pollution can increase allostatic load and activate the hypothalamic-pituitary-adrenal (HPA) axis in a manner similar to psychological stress (for review see Thomson, 2019); further highlighting the need for inclusive and representative participants samples, as individuals from different backgrounds might not demonstrate the same physiological responses to air pollution. Genetic risk for AD also moderated relationships between air pollution exposure and brain outcomes in two studies (Alemany et al., 2018; Essers et al., 2023). These findings are particularly interesting because AD incidence increases with air pollution exposure (for review see Hussain et al., 2023) and postmortem studies of children and young adults have identified AD-like pathologies in individuals who lived in highly polluted areas (Calderón-Garcidueñas et al., 2004). Finally, while several studies investigated sex differences in relationships between air pollution and brain outcomes, most studies did not find evidence of significant sex differences. Yet, previous studies of younger children have suggested that male children may be more susceptible to air pollution, while studies of older children demonstrate that female children may be more susceptible, perhaps due to developmental differences in lung size growth and hormonal maturation (Clougherty, 2010). Moreover, evidence from animal studies suggests several mechanisms of sex differences in exposure-related brain outcomes, potentially through differences in sex hormones and inflammation (see Cory-Slechta et al., 2023 for review). Future studies need to continue to explore psychosocial, environmental, and genetic factors that moderate these relationships, as research of this nature is vital to understanding who might be most vulnerable to the neurotoxic effects of air pollution.

### 4.3 Pollutant, Mixtures, Sources, and Geographical Considerations

Most studies examined PM_2.5_ or chemicals and components found within PM_2.5_. Far fewer studies examined gaseous pollutants. Importantly, many studies conducted various single and multiple pollutant models to try and identify differential effects of various exposures on the brain by accounting for potential confounding or co-exposures of other known environmental neurotoxins. Yet, given collinearity between certain pollutant types, determining unique effects of each pollutant on the brain may be difficult when participants are exposed to a mixture of pollutants. Moving forward, investigating various source(s) and chemical composition of PM exposure may help clarify potential discrepancies of findings between studies (Hamra and Buckley, 2018). Within the studied countries, the United States is much larger geographically compared to the other European nations sampled, and primary source(s) and composition of PM likely differ. For instance, over 50% of passenger vehicles in Europe are powered by diesel engines (European Environment Agency, 2022), whereas in the US the majority (>95%) of passenger cars and light trucks are fueled by gasoline engines (US Department of Energy, 2022). Moreover, the U.S. has greater reliance on gas-powered vehicles for transportation needs of United States residents compared with greater reliance on public transportation by European residents (Bertaud and Richardson, 2017). As such, additional studies examining PM components and considering how mixtures may impact brain structure and function are necessary to further clarify this emerging field and to help inform policy.

### 4.4 Confounders and Covariate Selection

While analytic models in these studies included some common variables (i.e., age at MRI, sex assigned at birth, race and/or ethnicity (or parental country of origin), intracranial volume for structural outcomes), there was considerable variability in specific sets of covariates used. Almost all studies included socio-demographic measures in their models; albeit many included individual-or family-specific measures (i.e., parental education and/or household income) whereas others included measures of community-or census tract-level measures (e.g., neighborhood SES). However, justification and inclusion of other potential covariates as confounders were less clear, posing a potential barrier to synthesizing results and assessing potential confounding bias across studies. Careful selection of covariates is critical for mitigating confounding bias in statistical analyses (Walter and Tiemeier, 2009). This issue is particularly pertinent when considering socioeconomic and demographic covariates–which reflect a host of social and structural factors (Cardenas-Iniguez and Gonzalez, 2024)–as air pollution exposure is related to both economic and racial inequality, especially in the United States (Brekel et al., 2024; Hajat et al., 2021, 2015; Rentschler and Leonova, 2023). Most studies, however, did not clearly articulate the rationale and conceptual framework for choosing each potential confounder and covariate of interest. While inclusion of these essential confounders is necessary to understand the independent contribution of air pollution exposure on the brain, studies that are overly inclusive of covariates may create multi-collinearity issues and/or over adjust for factors that may be on the causal pathway. Thus, researchers should provide strong rationale for inclusion of additional variables as well as take regional and cultural considerations into account to ensure their models include the appropriate variables to remove confounding effects specific to their study sample and their statistical modeling techniques.

### 4.5 Continuing Challenges

Each study reviewed takes an epidemiological approach to investigate relationships between air pollution exposure and brain features. This approach allows for a naturalistic observation of how everyday exposure to a range of pollutants impacts the brain across heterogeneous populations and geographic regions. While epidemiological studies offer the possibility of identifying vulnerable periods of development, epidemiological and observational approaches cannot determine causality; rather we must compare brain phenotypes across participants with varying levels of air pollution exposure. In addition, the application of geospatial exposure models to large cohort studies comes with its own set of strengths and limitations. These complex spatial temporal models can be applied on a large scale to provide a precise and continuous map of pollutant exposure across broad geographic regions; on the other hand, measurement errors and data missingness pose challenges to model accuracy (Cui et al., 2022). Furthermore, it is difficult to tease apart relative contributions of different pollutants given the inevitability of co-exposure in nature. Thus, animal models may shed more light on mechanisms of action. Epidemiological studies should continue working alongside animal models to understand the neural mechanisms underlying observed associations between air pollution exposure and brain development (for review see Costa et al., 2020; Zundel et al., 2022).

On the outcome side, participants must remain still in the scanner over extended periods of time for MRI to be usable. Poor image quality and missing data arise from an inability to complete the scanning session and head motion in the scanner in pediatric populations. As children who are younger, of lower socioeconomic status, and suffer from neurodevelopmental disorders (e.g., ADHD) tend to move more in the MRI scanner, this data loss is not equally distributed across sample demographics (Cosgrove et al., 2022). While statistical methods can partially correct this selection bias, this issue limits generalizability of findings.

Regardless of these challenges, with 99% of the world still living in areas with air pollution levels known to cause harm to human health (WHO, 2024), this emerging body of research suggests there is a rightful concern for the long-term health effects that chronic air pollution exposure might have on the developing brain. Alongside the call for additional research, this growing field of research supports recent public policy changes (i.e., California Children’s Right to Healthy Brain Development) that protect and ensure child brain health.

## 5. Conclusions

Outdoor air pollution exposure during critical windows of brain development such as pregnancy, childhood, and early adolescence has been associated with widespread structural and functional brain outcomes, albeit with mixed associations across studies. The literature has grown considerably in the past 5 years, yet there are still few longitudinal studies and key questions regarding the potential sensitive period(s) of exposure on brain development remain to be determined. Moving forward, the developmental periods and neural mechanisms with greatest susceptibility remain to be determined, as do their potential lifespan consequences for cognition, behavior, and mental health.

## Data Availability

No data were used in this systematic review.

## Acknowledgments

Research described in this article was supported by the National Institutes of Health (MMH: NIEHS R01ES032295, R01ES031074; KLB: P30ES07048-27, K99MH135075; JMM: T32ES013678; MAR: 8K00ES036895-03)

## Author Contributions

Conceptualization: MMH, JM, MD

Data curation: JM, MD, MAR, DLC, KLB

Formal Analysis: JM, MD, MAR

Funding acquisition: MMH

Methodology: MMH, JM, MD

Project administration: MMH, JM, MD

Resources: MMH

Software:

Supervision: MMH

Visualization: DLC, MAR, KLB

Writing – original draft: JM, MD

Writing – review & editing: MMH, DLC, MAR, KLB

