## Supplement for "A Systematic Review of Air Pollution Exposure and Brain Structure and Function during Development"

**Supplemental Material**

| **Brain / Imaging Terms**  Magnetic resonance image(s)  Magnetic resonance imaging  MRI  White matter hyperintensity  White matter hyperintensities  Neuroimage(s)  Neuroimaging  Neuroinflammation  Systemic inflammation  White matter volume(s)  Brain structure  Brain volume(s)  Neurotoxic  Neurotoxicity  Neurotoxicities  Functional connectivity  Brain/pathology  Brain/physiopathology  Cognition Disorders/pathology  Cognition Disorders/chemically induced  White Matter/pathology | **Air Pollution Terms**  Air Pollution  Air Pollutant(s)  Particulate Matter  Ozone  Nitrogen dioxide(s)  Nitrogen oxides  Sulfur Dioxide  Black carbon  Elemental carbon  Vehicle Emission(s)  Diesel  Diesel exhaust(s)  Vehicle exhaust(s)  Vehicular exhaust(s)  Road traffic  PM_2.5_  PM_10_  Coarse particle(s)  Coarse particles  Ultrafine particle(s)  Polycyclic aromatic hydrocarbon(s)  Air Pollution/adverse effects  Polycyclic Aromatic Hydrocarbons/adverse effects  Polycyclic Aromatic Hydrocarbons/poisoning  Polycyclic Aromatic Hydrocarbons/toxicity  Inhalation Exposure/adverse effects |
| --- | --- |

**Supplemental Table 1**. **Table of search terms used in literature search.** Combinations of exposure and outcomes terms used to identify relevant articles.

| **Study (Author/Year)** | **Scanner and Modality Details** | **Preprocessing** |
| --- | --- | --- |
| Peterson et al. 2015 | 3T GE with 8-channel head coil; T1 weighted | ANALYZE 8.0 and in-house software; structural MRI's, surface mapping |
| Pujol et al. 2016a (NeuroImage) | 1.5T GE with 8-channel head coil; T1 weighted; diffusion weighted (25 dir.; b0; b1000); rs-fMRI (eyes closed); Sensory task fMRI; MRS | VBM and FREESURFER (version not reported); DTI via FMRIB FDT; SPM (version 8) and functional connectivity maps using 3.5 mm spheres of frontal lobe (n=4) and caudate (n=2) seeds; |
| Pujol et al. 2016a (Brain and Behavior) | 1.5T GE with 8-channel head coil; T1 weighted; diffusion weighted (25 dir.; b0; b1000); rs-fMRI (eyes closed) | VBM and FREESURFER (version not reported); DTI via FMRIB FDT; SPM (version 8) and functional connectivity maps using 3.5 mm spheres of frontal lobe (n=4) and caudate (n=2) seeds; |
| Mortamais et al. 2017 | 1.5T GE with 8-channel head coil; T1 weighted | FREESURFER volumes (version not reported) |
| Alemeny et al. 2018 | 1.5T GE with 8-channel head coil; T1 weighted | FREESURFER (version 5.3) |
| Guxens et al. 2018 | 3T GE with 8-channel head coil; T1 weighted | FREESURFER (version 5.1) |
| Brunst et al. 2019 | 3T Philips Achieva with 32-channel head coil; T1 weighted; MRS | LCModel commercial software; adjusted for the tissue contributions from gray matter, white matter and cerebrospinal fluid (CSF) using FSL; adjusted to the T1 and T2 relaxation decay rate corrected water concentration, and  corrected for literature reported T1 and T2 relaxation decay rates of the primary metabolites. |
| Beckwidth et al. 2020 | 3T Philips Achieva with 32-channel head coil; T1 weighted | SPM12 and FREESURFER (version not reported) |
| Cserbik et al. 2020 | 3T scanners (Siemens Prisma, GE 750, Philips) with 32- or 64-channel head coil; T1 weighted | ABCD pipeline – FREESURFER (version 5.3) |
| Burnor et al. 2021 | 3T scanners (Siemens Prisma, GE 750, Philips) with 32- or 64-channel head coil; T1 weighted; diffusion weighted (96-dir., b0; b500; b=1000; b=2000; b=3000) | ABCD pipeline; AtlasTrak white matter tractography; FA/MD; RSI |
| Lubczynska et al. 2021 | 3T GE with 8-channel head coil; T1 weighted | FREESURFER (version 6.0) |
| Binter et al. 2022 | 3T GE with 8-channel head coil; T1-weighted and diffusion weighted | FREESURFER (version 6.0) |
| Bos et al. 2022 | 3T Philips Achieva with 32-channel head coil; T2 weighted | in-house neonatal-specific automated pipeline |
| Miller et al. 2022 | 3T GE with 32-channel head coil; T1 weighted | Tensor based morphometry via ANTs and Jacobian determinant image |
| Perez-Crespo et al. 2022 | 3T GE with 8-channel head coil; T1-weighted; rs-fMRI (eyes closed) | 5 min and 52s of resting-state fMRI collected with eyes closed. fMRIPrep software; de-spiking was applied, and the cerebrospinal fluid, white matter and global signals, as well as motion parameters (and their quadratic terms and temporal derivatives) were regressed out of the data. Pairwise correlation coefficients amongst 380 brain areas were grouped into 31 regions based on location and common properties and then into 5 different brain functional networks: auditory, somato-sensory/motor, visual, task positive, and task negative, and a 6th group comprising subcortical structures and the cerebellum. |
| Peterson et al. 2022 | 3T GE with 8-channel head coil; T1 weighted; diffusion weighted (15 dir, b0; b1000); ASL (PASL); MRS | ANALYZE 7.5 software; BrainSuite; in-house software for deformation-based morphometry; DSI Studio |
| Cotter et al. 2023 | 3T scanners (Siemens Prisma, GE 750, Philips) with 32- or 64-channel head coil; T1 weighted & rs-fMRI (eyes open) | ABCD pipeline: Resting-state data was collected in two sets of two five-minute acquisition periods, for a total of twenty cumulative minutes, to increase the likelihood of 12.5 minutes of low-motion (framewise displacement < 0.2 mm) available data. Subjects were instructed to keep their eyes open and fixed on a crosshair. Intrinsic brain networks were defined using Gordon’s network parcellation. |
| Essers et al. 2023 | 3T GE with 8-channel head coil; T1-weighted | FREESURFER (version 6.0) |
| Sukumaran et al. 2023 | 3T scanners (Siemens Prisma, GE 750, Philips) with 32- or 64-channel head coil; T1 weighted; diffusion weighted (96-dir., b0; b500; b=1000; b=2000; b=3000) | ABCD pipeline; FREESURFER (version 5.3); RSI |

**Supplemental Table 2**. **Details about magnetic resonance imaging (MRI) scanners and preprocessing pipelines used for brain image acquisition.** Abbreviations: ABCD = Adolescent Brain Cognitive Development Study; ANTs = Advanced Normalization Tools; ASL = Arterial Spin Labeling; DTI = Diffusion Tensor Imaging; GE = General Electric; MRS = Magnetic Resonance Spectroscopy; rs-fMRI = resting state functional MRI; RSI = restriction spectrum imaging; SPM = Statistical Parametric Mapping; T = Tesla; VBM = Voxel-Based Morphometry.

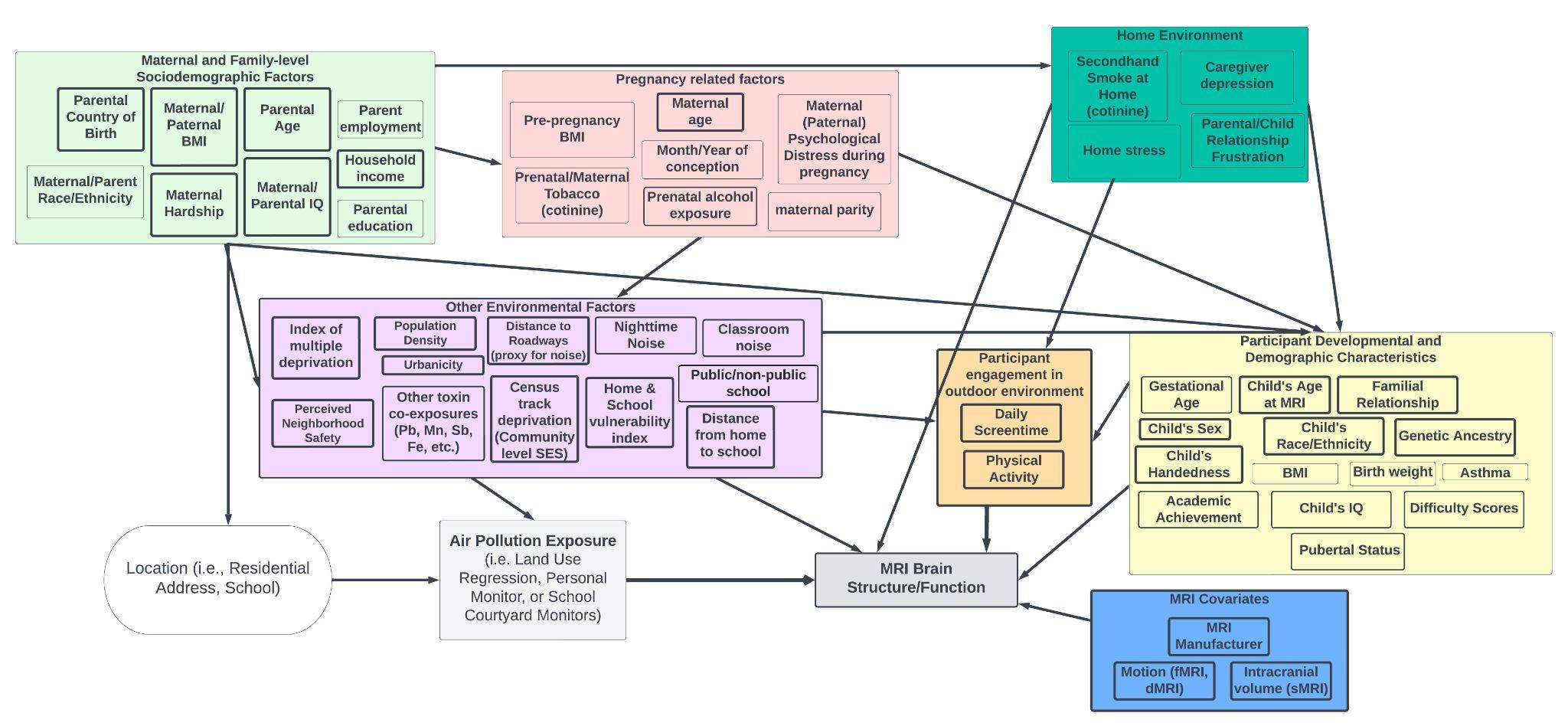

**Supplement Figure 1. Conceptual, simplified directed acyclic graph (DAG) and content knowledge used to consider potential confounders in studying outdoor air pollution and brain development.** Set of variables considered as potential confounders between air pollution exposure (i.e. independent variable, light gray) and brain MRI (i.e., dependent variable, dark gray) are grouped into maternal and family-level socioeconomic factors, pregnancy related factors, home environment, other environmental co-exposure factors, participant engagement with outdoor environment, participant developmental and demographic factors, and essential MRI covariates. Most studies are based on outdoor air pollution exposure at residential address or school courtyard (i.e., white oval), so upstream and downstream factors of living/schooling in a given location are considered in that context. Of note, the variables listed in each domain are directly taken from the included 20 studies and grouped into their respective categories; albeit many studies did not include a DAG and/or a rationale for conceptualizing confounders. In fact, many participant demographics and developmental characteristics adjusted for could conceivably also fall along the causal pathway between air pollution and MRI outcomes (i.e. physical, cognitive, and mental health factors of the child, i.e. BMI, birth weight, asthma, IQ, etc.).

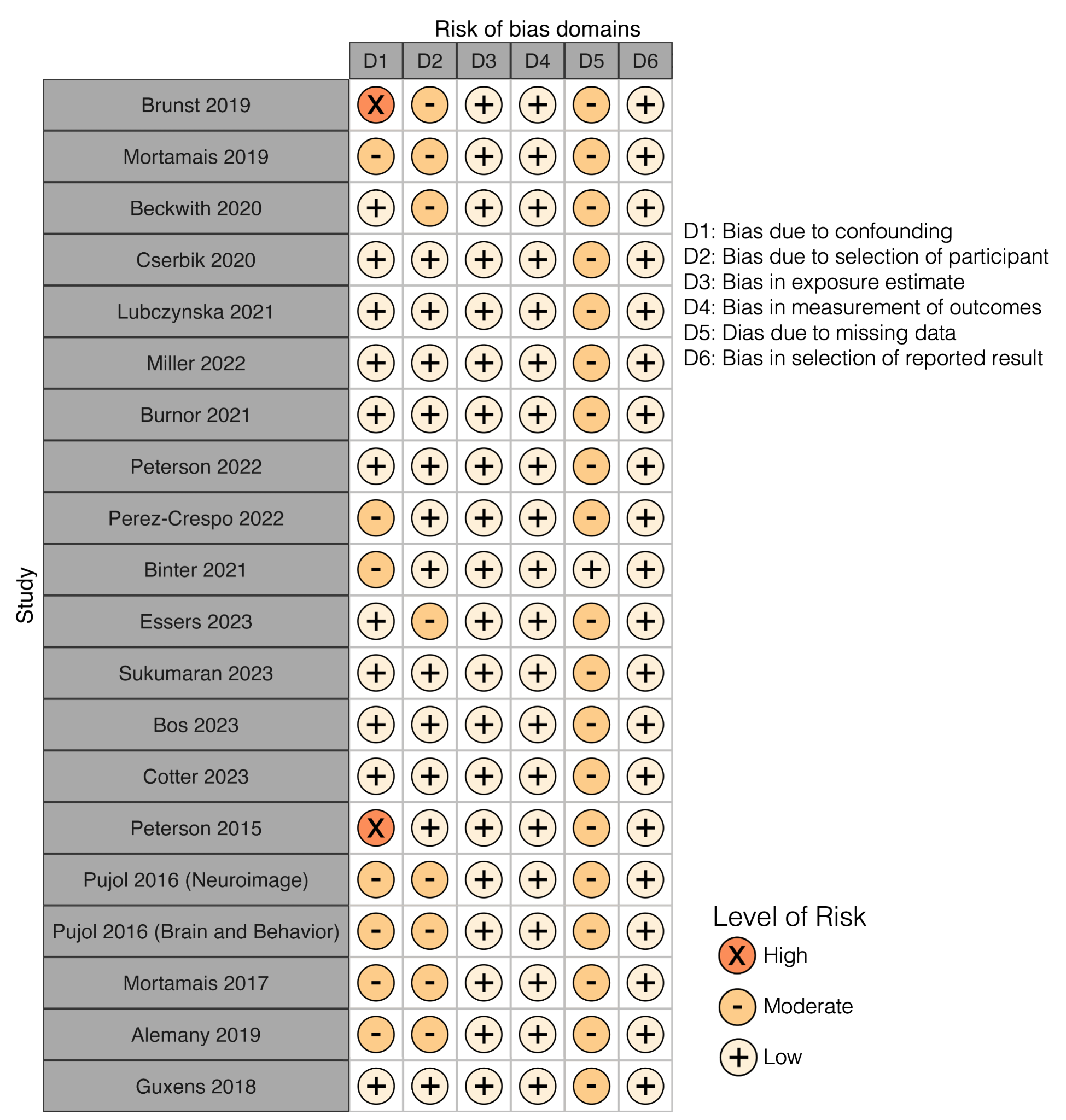

**Supplemental Figure 2. Risk of bias scores for each domain per study included (n=20) on outdoor air pollution and brain MRI outcomes in children and adolescents.**
